# Convergent human genetic evidence implicates serine biosynthesis in diabetic peripheral neuropathy

**DOI:** 10.64898/2026.06.09.26355286

**Authors:** Vera Fridman, Aastha Kakar, Aubrey Jensen, Liedewei Van de Vondel, Allison Wheeler, Lawrence S. Phillips, Jin Zhou, Stephan Zuchner, Jane Reusch, Sridharan Raghavan

**Affiliations:** Department of Neurology, University of Colorado Anschutz Medical Campus, Aurora, CO; Department of Biomedical Informatics, University of Colorado Anschutz Medical Campus, Aurora, CO; Phoenix Veterans Affairs Medical Center, Phoenix, AZ; Department of Biostatistics, UCLA Fielding School of Public Health, Los Angeles, CA; Dr. John T. Macdonald Foundation Department of Human Genetics and John P. Hussman Institute for Human Genomics, University of Miami Miller School of Medicine, Miami, FL; Atlanta Veterans Affairs Medical Center, Decatur, GA; Department of Medicine, Emory University, Atlanta, GA; Medicine Service, US Department of Veterans Affairs Eastern Colorado Health Care System, Aurora, CO; Division of Endocrinology, Diabetes, and Metabolism, Department of Medicine, University of Colorado Anschutz Medical Campus, Aurora, CO; Section of Academic Primary Care, US Department of Veterans Affairs Eastern Colorado Health Care System, Aurora, CO; Division of General Internal Medicine, University of Colorado Anschutz Medical Campus, Aurora, CO

**Keywords:** Neuropathy, diabetic peripheral neuropathy, genome wide association study, serine

## Abstract

**Background:** Diabetic peripheral neuropathy (DPN) is a common and disabling complication of diabetes for which no disease-modifying therapies are currently available. Glycemic and metabolic drivers do not fully explain why only a subset of individuals with diabetes develop DPN, and underling genetic contributors remain poorly defined.

**Methods:** We performed a multi-population GWAS of neuropathy in individuals with and without diabetes using the VA Million Veteran Program and UK Biobank, with replication in the All of Us Research Program (AoU). Gene-based and gene-set analyses were used to identify enriched biological pathways. The relationship between circulating serine levels and DPN was further investigated, using two-sample Mendelian randomization. To extend the findings beyond common variation and DPN, we assessed the burden of rare, predicted high-impact variants in GWAS-prioritized genes in individuals with unsolved inherited neuropathies using the GENESIS platform.

**Findings:** Among individuals with type 2 diabetes, we identified seven genome-wide significant loci (p<5×10□ □) including variants in *PHGDH* and *PSPH*, key enzymes in serine biosynthesis, *TEAD1, CYP4F11, LARGE1, FTO*, and *COBLL1*. No significant loci were identified in individuals without diabetes or with type 1 diabetes. Four loci (*PHGDH, TEAD1, FTO* and *CYP4F11)* replicated in AoU (*p* <0·05). Mendelian randomization showed that higher genetically predicted serine levels were associated with lower DPN risk, consistent with a causal role of serine metabolism in disease pathogenesis. Rare-variant burden analyses demonstrated association of predicted deleterious variants with inherited neuropathy cases for *PHGDH* (odds ratio [OR] 12.7 [95% CI 7·9, 20·4]), *PSPH* (OR 8·5 [7·2, 10·2]), *PHKG1* (OR 4·8 [3·7, 6·3], and *LARGE1* (OR 0·007 [0·0004, 0·1]).

**Interpretation:** Convergent genetic evidence across common and rare variation implicates the serine synthesis pathway in DPN susceptibility. These findings link diabetic and inherited neuropathies through a shared metabolic mechanism and provide a genetically supported rationale for investigating serine-directed therapeutic strategies.

**Funding:** This research is based on data from the Million Veteran Program, Office of Research and Development, Veterans Health Administration, and was supported by MVP000 and by awards MVP009/MVP037 I01-BX005831 and MVP051. S.R. is supported by VA award I01-BX006417. V.F. is supported by NIH award 1K23DK118202-01A1. S.Z. is supported by the CMT Association, CMT Research Foundation, All of US Research Program (3OT2OD037907), and NIH (1R21HG013397, 5R01NS072248). J.R. is supported by VA awards BX002046 and CX001532, NIH awards DK124344, HL165433, AG087809, P30DK116073, UM1 TR004399, and the Ludeman Center. L.S.P. is supported in part by VA awards CSP #2008, I01 CX001899, I01 CX001737, and I01 BX005831, NIH awards R01 DK127083, R03 AI133172, R21 AI156161, U01 DK098246, UL1 TR002378, and a Cystic Fibrosis Foundation award PHILLI12A0. L.VdV. was supported by the Peripheral Nerve Society Laura Feltri Basic Research Fellowship.

## Introduction

Diabetes is the most common cause of neuropathy worldwide, and in 2021, diabetic peripheral neuropathy (DPN) ranked among the top five neurological contributors to global fatal and non-fatal disease burden.^1^ Symptoms of DPN include sensory loss, neuropathic pain, gait difficulty, and a propensity to limb-threatening foot ulcers. Beyond its profound impact on daily function and quality of life, DPN is associated with a substantially increased risk of all-cause mortality.^2^ Despite rapidly evolving insights into the biology of DPN, no disease modifying treatments beyond optimization of glycemic and metabolic risk factors are currently available.

A critical question in the neuropathy field has been why only a subset of individuals develop DPN, while others are spared nerve injury. Prior genome-wide association studies (GWAS) of DPN have revealed several risk loci with potential relevance to neuropathy and neuropathic pain, however, have not highlighted therapeutically targetable pathways.^3–6^ The serine metabolic pathway is increasingly supported by genetic, metabolic, and clinical trial data to play a role in inherited neuropathies, diabetes, and DPN. These data raise the possibility that genetic variation in serine metabolism may be contributing to neuropathy development in people with diabetes, however prior GWAS have not linked serine with neuropathy.^7–12^

In this study, we leveraged the expansion of large-scale clinical-genomic biobanks to perform the largest GWAS of neuropathy to date in individuals with and without diabetes.^13–15^ We identified seven novel genome-wide-significant loci associated with neuropathy in type 2 diabetes and unmasked a new relationship between the therapeutically targetable serine biosynthetic pathway and DPN.

### Research in Context

#### Evidence before this study

Diabetic peripheral neuropathy (DPN) is one of the most common and disabling complications of diabetes and is a major cause of morbidity worldwide. Despite its prevalence, no disease modifying treatments currently exist and the biological mechanisms underlying individual susceptibility to DPN remain incompletely understood. We searched PubMed for articles published in English up to June 15, 2025, using the terms “diabetic neuropathy”, “diabetic peripheral neuropathy/DPN”, “neuropathy”, “genome-wide association study/GWAS”, “serine”, “*PHGDH”*, and *“PSPH”*. Prior GWAS have identified risk loci with potential relevance to DPN and neuropathic pain but have been limited by sample size and heterogeneous case definitions (case sample sizes 572-40,475). Alterations in serine and sphingolipid metabolism have been implicated in diabetic and inherited neuropathies, and genetic variation in serine biosynthesis has been associated with rare neuropathic conditions but has not been linked to DPN.

#### Added value of this study

This is the largest GWAS of neuropathy to date (more than one million participants across discovery and replication cohorts) and is the first to identify an association of variants in serine synthesis genes (*PHGDH* and *PSPH)* with DPN. Follow up gene-based analyses, pathway enrichment, Mendelian randomization and rare-variant burden analyses in an independent unsolved inherited neuropathy cohort independently supported a role for serine synthesis in DPN susceptibility. Taken together, these findings suggest that DPN and inherited neuropathies may share an underlying genetic and metabolic vulnerability in the serine biosynthetic pathway.

#### Implications of all the available evidence

The role of serine metabolism in neurological disease has previously been underscored in hereditary neuropathies, and both preclinical and clinical studies have demonstrated that the serine metabolic pathway can be targeted therapeutically. The identification of common DPN risk loci in serine synthesis genes, together with pre-existing metabolic and experimental data, link DPN to rare inherited neuropathies through a potential shared metabolic mechanism. Given the targetable nature of the serine and sphingolipid pathways, the current findings could offer a foundation for novel treatment strategies for DPN.

## Methods

### Data sources and study participants

Biobank data for this study came from three previously described cohorts: the Million Veteran Program (MVP), the UK Biobank, and the All of Us Research Program (AoU).^13,16,17^. Ethical and human subjects research oversight for the MVP and this specific project were provided by the VA Central Institutional Review Board. We stratified the samples into three mutually exclusive cohorts: those with type 2 diabetes, those with type 1 diabetes, and those without diabetes. AoU served as a replication sample, and only individuals with type 2 diabetes were included for replication of GWAS findings. Details of how individuals with and without diabetes were identified are provided in the Supplemental Methods.

Genotyping, genotyping quality control, and imputation in the MVP, UK Biobank, and AoU have been previously described^13–16^. For MVP and UK Biobank, genotyping of DNA extracted from blood was performed on a customized Affymetrix Axiom biobank array (MVP 1·0 Genotyping Array) and the UK Biobank Axiom array, respectively. For AoU, we examined short-read whole genome sequencing (WGS) data from the AoU controlled tier dataset V8.^15^ Additional details regarding sample and genotyping quality control for each data source are provided in the Supplemental Methods.

### Neuropathy case and control definitions

Neuropathy cases in MVP were defined by the presence of at least one occurrence of a phecode (an aggregation of related diagnosis codes) for a neuropathy condition (Supplemental Table 1).^18,19^ Controls were those who did not meet the case definition in each cohort. As a sensitivity analysis, we repeated the GWAS using a stringent case definition that required at least two uses of a neuropathy-related phecode.

Neuropathy cases in UKB were defined based on a combination of primary care, ICD-9, and ICD-10 diagnosis codes for neuropathy and neuropathic conditions (Supplemental Table 2). Controls were individuals who did not meet the case definition in each cohort. As in MVP, we performed a sensitivity analysis requiring at least two uses of the case-defining codes for classification as a case. We performed an additional sensitivity analysis excluding individuals with hereditary neuropathies among the cases. In AoU, neuropathy cases were defined based on the same set of ICD codes mapped to the phecodes used in MVP (Supplemental Table 1).

### Statistical analysis

#### GWAS

We performed race/ethnicity stratified GWAS of autosomes followed by multi-population meta-analysis in three mutually exclusive cohorts based on diabetes status – those with type 1 diabetes, type 2 diabetes, and without diabetes. In MVP, we used the Harmonized Ancestry and Race/Ethnicity (HARE) algorithm that integrates genetic and self-reported race/ethnicity information to assign individuals into one of four genetically-informed race/ethnicity groups: Non-Hispanic White (NHW), Non-Hispanic Black (NHB), Hispanic (HIS), and Asian Americans.^20^ Due to sample size limitations in specific populations, we limited analysis in UKB to individuals of European ancestry, defined by both self-reported ethnic background and PCs, and in individuals with type 1 diabetes in MVP to NHW, NHB, and HIS. Within each population group, imputed and directly measured genetic variants were tested for association with neuropathy case status using logistic regression assuming an additive genetic model adjusted for age at enrollment (specified as decade categories in MVP), sex, and the first ten population-specific genetic principal components (PCs) using SAIGE, a random effects modeling software that accounts for sample relatedness.^21^ Population-specific GWAS results from MVP and UKB were combined using inverse variance weighted fixed-effects meta-analysis in METAL.^22^ For replication in individuals with type 2 diabetes in AoU, the top SNPs associated with neuropathy in the primary multi-population GWAS meta-analysis were tested for association with neuropathy case status in an additive genetic model adjusted for age, sex, and ten principal components, stratified by genetic ancestry into three mutually exclusive groups: European ancestry (EUR), African ancestry (AFR), and Amerindian ancestry (AMR). We considered *p* <0·05 to be suggestive of replication in AoU.

To summarize the GWAS results, we used FUMA (http://fuma.ctglab.nl/), a platform that annotates, prioritizes, and visualizes GWAS results,^23^ estimated the SNP-based heritability of DPN, and evaluated whether multiple, independent signals explained the associations at genome-wide significant loci (Supplemental Methods). To examine sensitivity of the GWAS to study design choices, we repeated the GWAS in individuals with type 2 diabetes using the more stringent case definitions described above and in models including body mass index (BMI) at MVP enrollment as a covariate. The latter model was included to assess independence of neuropathy genetic associations from adiposity, a strong risk factor for both neuropathy and diabetes.^24,25^

#### Follow-up analyses of GWAS results

Supplemental Figure 1 diagrams the analyses conducted as part of this study. Full description of analytic methods can be found in the Supplemental Methods. We used FUMA to search for expression quantitative trait locus (eQTL) associations of genome-wide-significant variants and to test for tissue-specific enrichment in expression of any genes mapped on the basis of physical position or eQTL associations.^23^ We used Multimarker Analysis of GenoMic Annotation (MAGMA) to perform gene-based association tests with neuropathy case status and gene set enrichment analysis (GSEA).^26^ We performed three genetic analyses to evaluate the relationship of serine with neuropathy: we estimated genetic correlation of plasma serine levels and DPN using LD Score Regression;^27^ we tested for statistical evidence that DPN and plasma serine levels share a causal genetic variant at two loci with genome-wide significant associations with both traits, *PHGDH* (chr 1) and *PSPH* (chr 7); we performed two-sample Mendelian randomization (MR) to test for evidence supporting a causal association of serine levels with DPN using genetic instruments for plasma serine levels drawn from six independent genetic studies in diverse populations. Finally, to help identify possible causal genes at the GW-significant loci and shared pathogenic mechanisms between DPN and inherited neuropathies, we examined burden of rare, potentially high-impact variants in 2099 individuals with unsolved inherited neuropathy in the GENESIS database^28^ compared to controls in AoU without neuropathy. In this analysis, we interrogated 11 genes highlighted in the GWAS results: *PHGDH, PSPH, SUMF2, CCT6A, PHKG1, CHCHD2, TEAD1, ARNTL, CYP4F2, CYP4F11*, and *LARGE1*. We used two strategies to prioritize rare, plausibly pathogenic variants – CADD PHRED score and Maverick score^29,30^ – and employed two genetic models, one similar to an additive model and a second evaluating a dominant effect of predicted high-impact variants.

## Results

### Multi-population GWAS of neuropathy identifies novel loci associated with DPN

The numbers of neuropathy cases and controls contributed by each cohort and population group are summarized in Table 1. The multi-population GWAS meta-analysis included a total of 693,804 participants without diabetes (46,461 cases), 287,832 with type 2 diabetes (132,470 cases), and 50,508 participants with type 1 diabetes (28,710 cases).

**Table 1.**
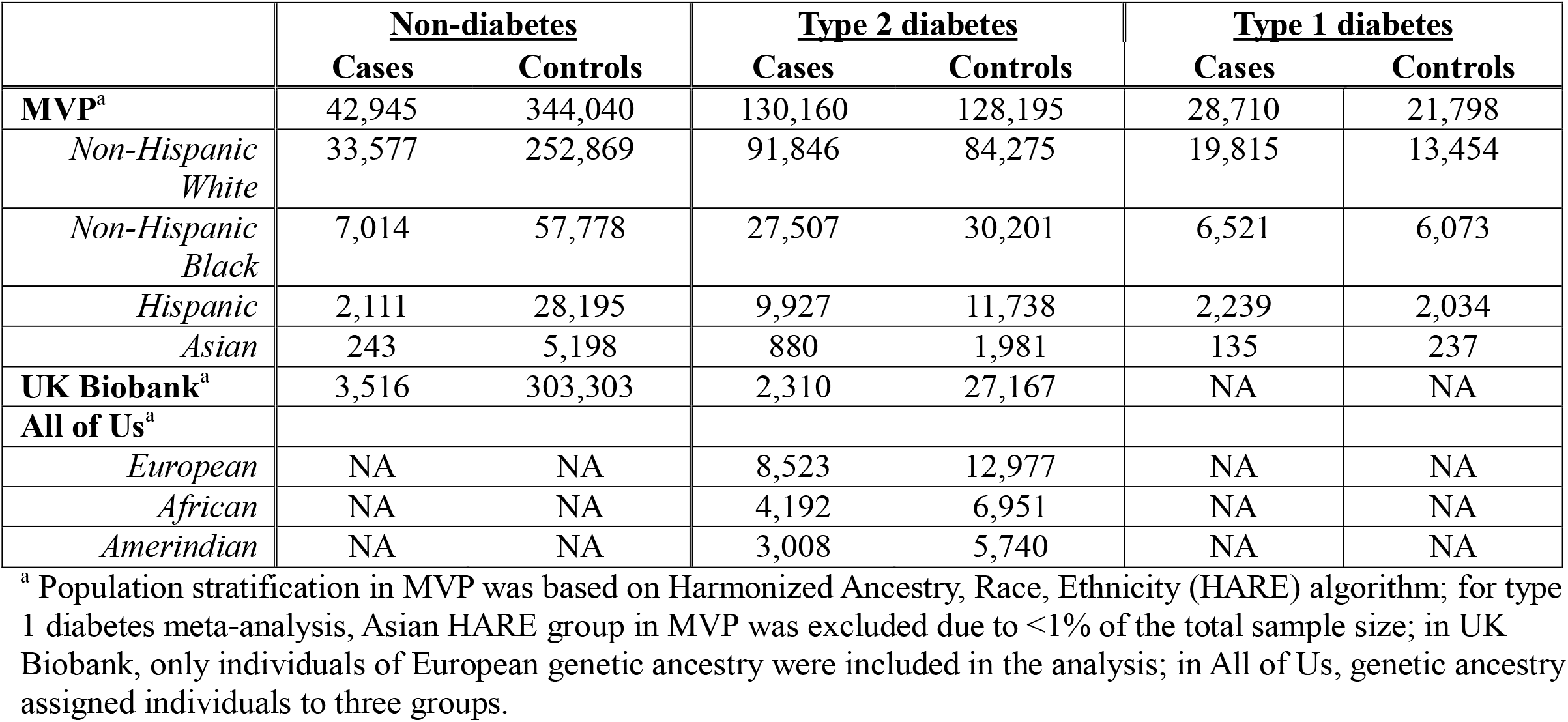
Study participant case/control and race/ethnicity or ancestry distribution.

Among individuals with type 2 diabetes the meta-analysis identified seven genome-wide significant loci associated with neuropathy (Figure 1A, Table 2, Supplemental Figure 2). These included rs562038, an intronic variant within *PHGDH* on chromosome 1, (effect allele frequency [EAF] 0·71, odds ratio [OR] 0·95 *p* = 4·1×10^−12^); rs3923113, a noncoding variant between *COBLL1* and *GRB14* on chromosome 2 (EAF 0·58, OR 1·04, *p* = 1·0×10^−8^); rs2016724, intronic variant within *PSPH* on chromosome 7, (EAF 0·52, OR 1·03, *p* = 8·8×10^−9^); rs2099744, intronic variant within *TEAD1* on chromosome 11 (EAF 0·63, OR 0·96, *p* = 1·2×10^−9^); rs62048402, intronic variant within *FTO* on chromosome 16 (EAF 0·38, OR 1·05, *p* = 2·2×10^−14^); rs11670222, intronic variant within *CYP4F11*on chromosome 19 (EAF 0·59, OR 1·04, *p* = 2·4×10^−8^); and rs13058212, intronic variant within *LARGE1* on chromosome 22 (EAF 0·002, OR 11·2, *p* = 2·4×10^−8^). Conditional and Joint (COJO) analyses suggested a single independent association signal at each locus (Supplemental Table 4). Notably, *PHGDH* and *PSPH* encode the first and third enzyme of the de novo serine biosynthesis pathway, respectively.

**Figure 1.**
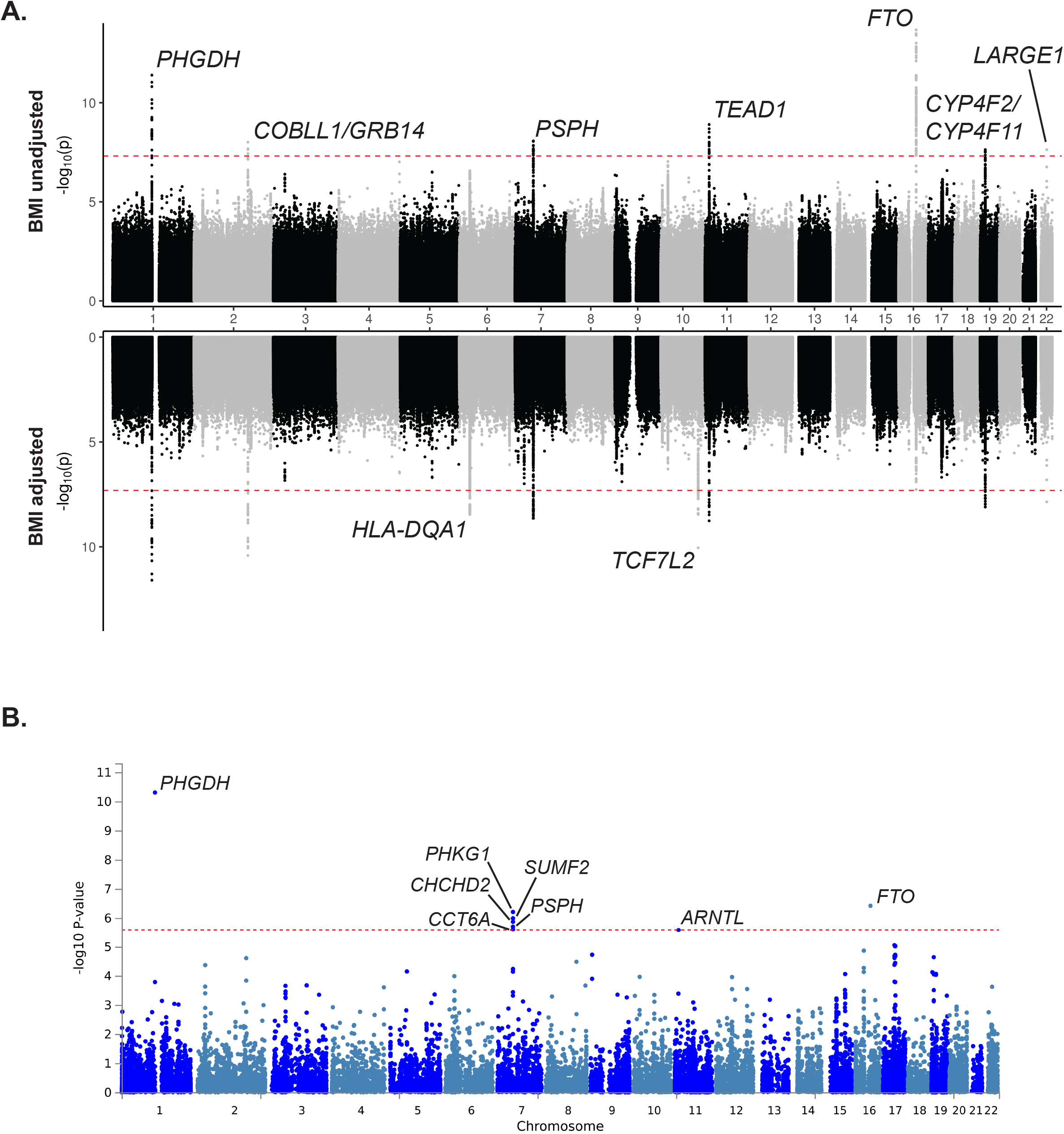
Miami plot showing results of multi-population genome-wide association meta-analysis of neuropathy in MVP and UK Biobank in models without adjustment for body mass index (BMI) (above the x-axis) and with adjustment for BMI (below the x-axis) in individuals with type 2 diabetes **(A).** Red dotted line indicates the threshold for genome-wide significance (*p* < 5×10^−8^). **(B)** Manhattan plot of gene-based analysis of multi-population genome-wide association study meta-analysis in individuals with type 2 diabetes identified seven genes with associations with neuropathy. Red dotted line indicates Bonferroni-corrected threshold for significance (*p* < 2·6×10^−6^ based on 19,283 tested gene associations). All labeled genes reached genome-wide significance except *ARNTL* which was just below the threshold for significance.

**Table 2.**
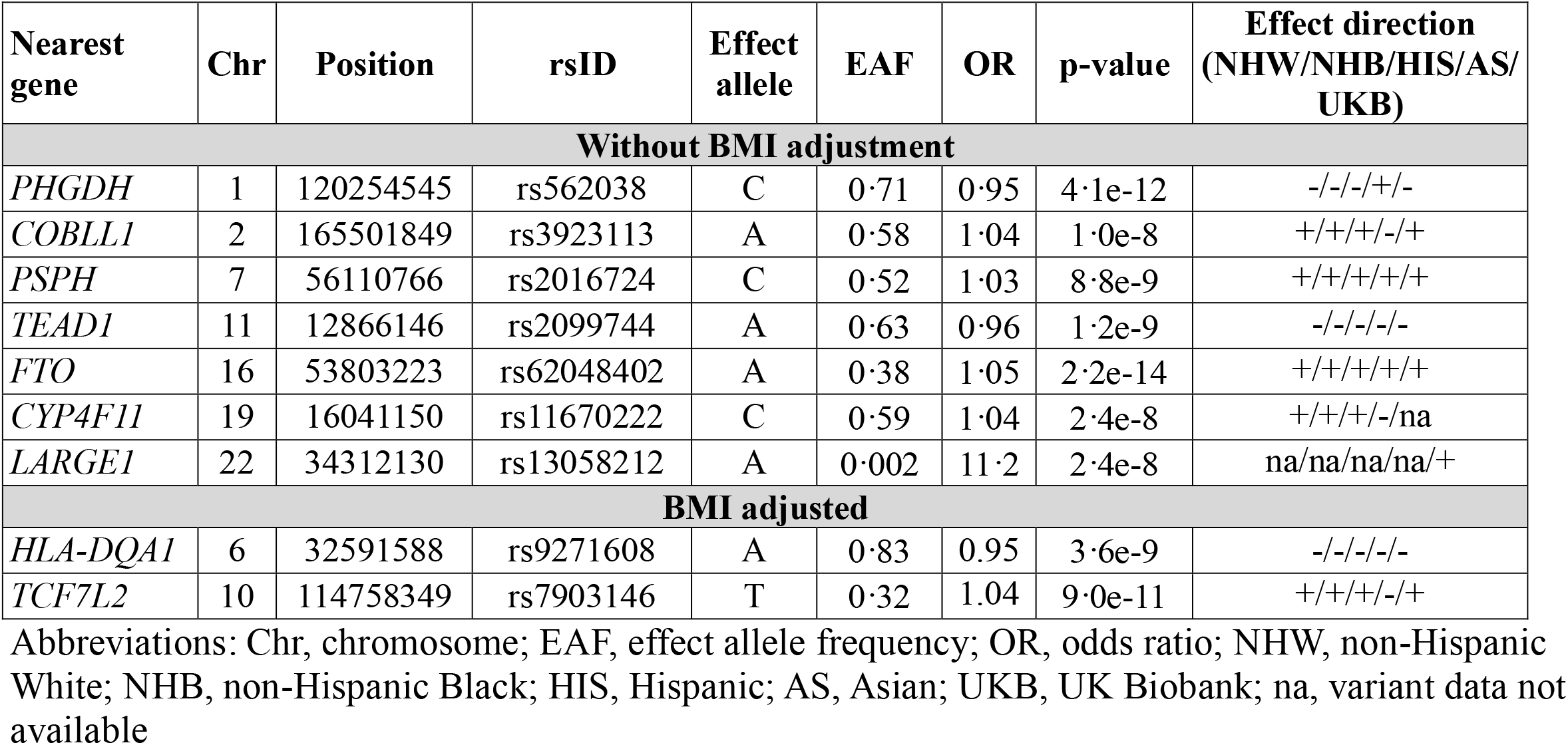
Top genome-wide significant associations with neuropathy in multi-ancestry meta-analysis in individuals with type 2 diabetes.

| Nearest gene | Chr | Position | rsID | Effect allele | EAF | OR | p-value | Effect direction (NHW/NHB/HIS/AS/UKB) |
| --- | --- | --- | --- | --- | --- | --- | --- | --- |
| **Without BMI adjustment** |  |  |  |  |  |  |  |  |
| *PHGDH* | 1 | 120254545 | rs562038 | C | 0.71 | 0.95 | 4.1e-12 | -/-/-/+/- |
| *COBLL1* | 2 | 165501849 | rs3923113 | A | 0.58 | 1.04 | 1.0e-8 | + /+ /+ /- /+ |
| *PSPH* | 7 | 56110766 | rs2016724 | C | 0.52 | 1.03 | 8.8e-9 | + /+ /+ /+ /+ |
| *TEAD1* | 11 | 12866146 | rs2099744 | A | 0.63 | 0.96 | 1.2e-9 | - /- /- /- /- |
| *FTO* | 16 | 53803223 | rs62048402 | A | 0.38 | 1.05 | 2.2e-14 | + /+ /+ /+ /+ |
| *CYP4F11* | 19 | 16041150 | rs11670222 | C | 0.59 | 1.04 | 2.4e-8 | + /+ /+ /- /na |
| *LARGE1* | 22 | 34312130 | rs13058212 | A | 0.002 | 11.2 | 2.4e-8 | na /na /na /na /+ |
| **BMI adjusted** |  |  |  |  |  |  |  |  |
| *HLA-DQA1* | 6 | 32591588 | rs9271608 | A | 0.83 | 0.95 | 3.6e-9 | - /- /- /- /- |
| *TCF7L2* | 10 | 114758349 | rs7903146 | T | 0.32 | 1.04 | 9.0e-11 | + /+ /+ /- /+ |

Abbreviations: Chr, chromosome; EAF, effect allele frequency; OR, odds ratio; NHW, non-Hispanic White; NHB, non-Hispanic Black; HIS, Hispanic; AS, Asian; UKB, UK Biobank; na, variant data not available

No genome-wide significant associations were identified in individuals without diabetes or those with type 1 diabetes (Supplemental Figure 3). Among the seven loci associated with neuropathy in type 2 diabetes, only rs562038 in *PHGDH* showed a nominal association with neuropathy in individuals without diabetes (Supplemental Table 5).

Adjustment for BMI identified two additional genome-wide significant loci: rs9271608, a noncoding variant located between *HLA-DRB1* and *HLA-DQA1* on chromosome 6 (EAF 0·83, OR 0·95, *p* = 3·6×10^−9^), and rs7903146, an intronic variant within *TCF7L2* on chromosome 10 (EAF 0·32, OR 1·04, *p* = 9·0×10^−11^) (Figure 1A, Table 2, Supplemental Table 6). In contrast, the *FTO* locus was no longer significant following BMI adjustment. All nine loci identified across the primary and BMI-adjusted analyses remained directionally consistent and statistically robust in sensitivity analyses using a more stringent neuropathy case definition (Supplementary Figure 4, Supplementary Table 7).

Replication analyses in the AoU cohort included 41,391 individuals with type 2 diabetes (15,723 cases) (Table 1). Among individuals of European ancestry, four of the seven loci identified in the primary GWAS replicated at nominal significance: rs562038 in *PHGDH* (EAF 0·68, OR 0·92, *p* = 5·0×10^−4^), rs2099744 in *TEAD1* (EAF 0·72, OR 0·94, *p* = 9·3×10^−3^), rs62048402 in *FTO* (EAF 0·44, OR 1·10, *p* = 2·6×10^−6^), and rs11670222 in *CYP4F11* (EAF 0·53, OR 1·05, *p* = 2·3×10^−2^). In additional ancestry groups, rs62048402 in *FTO* replicated among individuals of African ancestry (EAF 0·10, OR 1·11, *p* = 2·8×10^−2^) while rs2099744 in *TEAD1* replicated among individuals of admixed American ancestry (EAF 0·75, OR 0·93, *p* = 4·9×10^−2^) (Supplemental Table 8). Importantly, the direction of effect for all replicated loci in AoU was consistent with that observed in the discovery meta-analysis, supporting robustness and generalizability across independent cohorts.

We found no nominally significant associations for loci identified in eight published studies of neuropathic conditions in our multi-population GWAS analyses of individuals with and without diabetes (Supplemental Table 9).^3,4,6^ In contrast, five of 14 variants previously associated with macular telangiectasia demonstrated association with DPN including two variants in *PHGDH*, one in *PSPH*, and two in *CPS1* (Supplemental Table 10).

Using genomic location and eQTL information, FUMA mapped 25 genes to the 7 genome-wide significant loci (Supplemental Table 11). Notably, the lead *PHGDH* variant demonstrated a trans-eQTL association with *phosphoserine aminotransferase (PSAT1)* on chromosome 9, which encodes the second enzyme in the de novo serine biosynthesis pathway. Collectively, the 25 mapped genes showed significant enrichment for tissue-specific differential expression in skin and prostate tissues (Supplemental Figure 5).

Gene-based analysis of the multi-population GWAS summary statistics in individuals with diabetes identified seven genes associated with neuropathy at Bonferroni-corrected significance (*p* < 2·6×10^−6^; 19,283 genes tested). These included *PHGDH, FTO*, and five neighboring genes within the chromosome 7 locus – *PHKG1, SUMF2, PSPH*, CHCHD2, and *CCT6A* (Figure 1B).

### Convergent genetic evidence implicates serine biosynthesis in DPN

GSEA using Human Molecular Signatures Database^26,31^ identified two gene sets that achieved database-wide significance (*p* < 1·44×10^−6^ based on 34,775 gene sets tested): “Abnormal circulating serine concentration” from the Human Phenotype Ontology project^32^ (*p =* 3·9×10^−7^) and “G-protein coupled opioid receptor activity” from the Gene Ontology Molecular Function database^33^ (*p* = 8·9×10^−7^) (Supplemental Figure 6A). Analysis of five curated gene sets further demonstrated enrichment of GWAS signals in gene sets comprising genes previously implicated in neuropathy and serine biosynthesis (Set A), genes associated with macular telangiectasia and neuropathic conditions (Set B), and genes involved with glycine synthesis (Set C) (Supplemental Figure 6B, Supplemental Table 12). Together, the GWAS meta-analysis, regulatory analyses, and pathway enrichment reinforce serine biosynthesis as an important pathway underlying DPN susceptibility.

To further investigate the relationship between serine metabolism and neuropathy susceptibility several complementary genetic approaches were applied. First, plasma serine levels and neuropathy in individuals with type 2 diabetes demonstrated a genetic correlation of 46% based on GWAS summary statistics from individuals of European ancestry. Second, Bayesian colocalization analyses supported a shared genetic architecture for both traits at the *PHGDH* (posterior probability of shared causal variant 0·92) and *PSPH* (posterior probability of shared causal variant 0·72) loci (Supplemental Figure 7). Finally, two-sample MR analyses using genome-wide significant variants from six publicly available GWAS datasets of circulating serine levels representing diverse populations (Supplemental Table 13), consistently showed that higher genetically predicted serine levels were associated with lower odds of neuropathy (OR ranging from 0·85 [0·78 - 0·92] to 0·93 [0·89 - 0·98] per standard deviation increase in genetically predicted serine, all p<0·05) (Figure 2). The MR associations were consistent across multiple MR methods and sensitivity analyses (Supplemental Figure 8). Together these findings provide complementary genetic evidence supporting a relationship between serine metabolism and DPN susceptibility.

**Figure 2.**
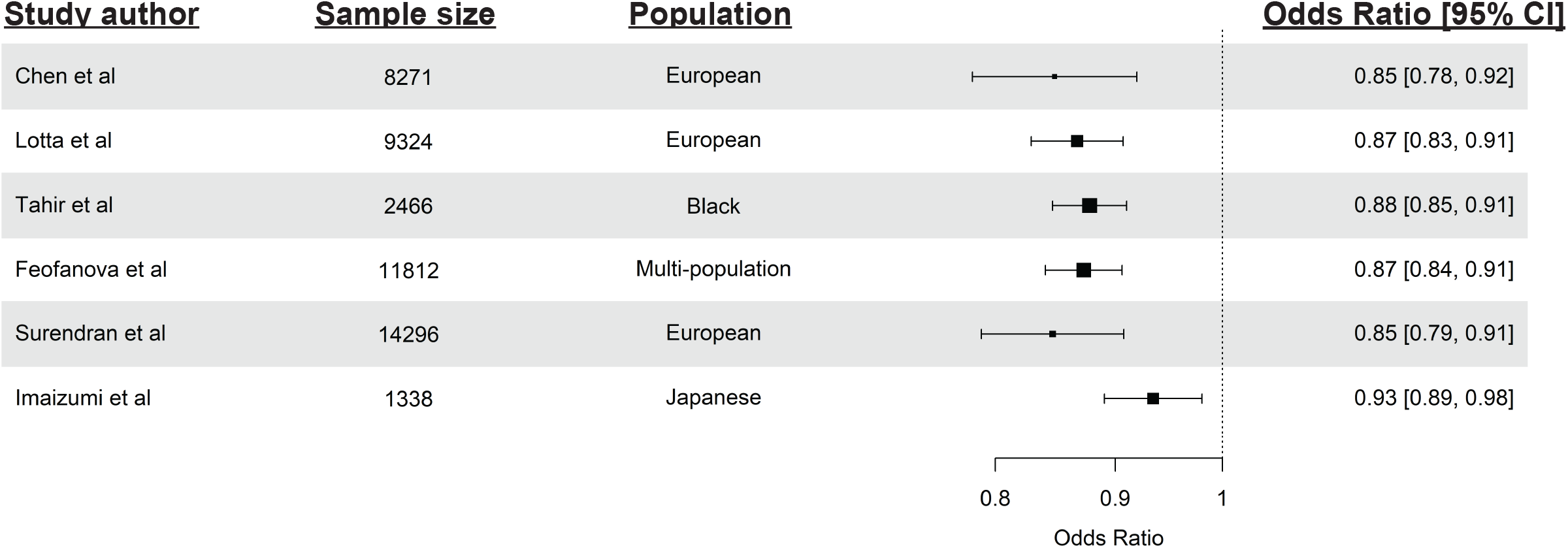
Two-sample Mendelian randomization analysis examining causal association of plasma serine levels with neuropathy in individuals with type 2 diabetes using genome-wide significant variants from six independent GWAS of plasma serine levels in diverse populations and summary statistics from multi-population GWAS meta-analysis of neuropathy in individuals with type 2 diabetes. Odds ratios shown indicate genetically-estimated neuropathy risk per genetically-estimated standard deviation increase in plasma serine levels.

### Rare, predicted high-impact variants in serine pathway genes are enriched in patients with inherited neuropathies

To determine whether the genetic signals identified in DPN extended beyond common variation, we examined an independent cohort with unresolved inherited neuropathies. Under an additive genetic model, rare variants predicted to have high functional impact were enriched in inherited neuropathy cases relative to controls in AoU using both variant prioritization algorithms for *PHGDH* (CADD OR 12·7 [95% CI 7·9, 20·4], Maverick OR 13·8 [10·5, 18·23]), *PSPH* (CADD OR 8·5 [7·2, 10·2], Maverick OR 18·4 [15·3, 22·1]), and *PHKG1* (CADD OR 4·8 [3·7, 6·3], Maverick OR 13·8 [10·5–18·2]) (Figure 3), and were depleted in neuropathy cases relative to controls in *LARGE1* (CADD OR 0·007 [0·00044, 0·11], Maverick OR 0·029 [0·0018, 0·47]) (Figure 3).

**Figure 3.**
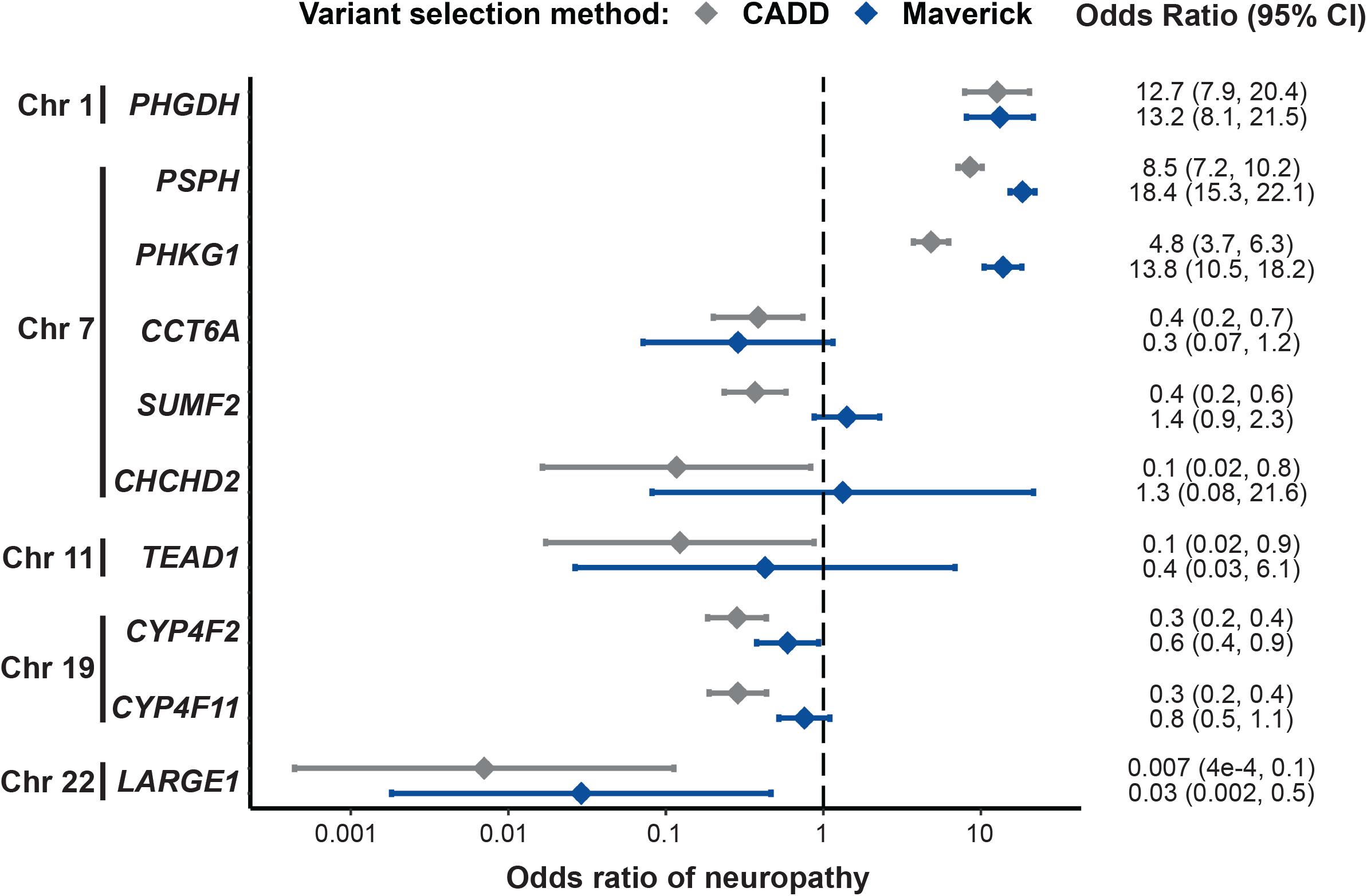
Gene-based burden testing of carriers of rare variants predicted to be of high functional impact based on two prediction algorithms (CADD and Maverick) in candidate genes identified in the GWAS, comparing individuals in the GENESIS registry of inherited neuropathies with control individuals in AllofUs. Odds ratios greater than one indicate enrichment of potentially high impact variants in these genes among individuals with inherited neuropathy relative to control individuals, suggesting that rare, high-impact variants in these genes may increase neuropathy risk. Odds ratios less than one indicate depletion of potentially high impact variants among individuals with inherited neuropathy relative to control individuals, suggesting rare, high-impact variants in these genes may decrease neuropathy risk.

Sensitivity analyses demonstrated that effect estimates generally moved further from unity as the Maverick algorithm was tuned to prioritize fewer variants with higher predicted functional impact (Supplemental Table 14). Under a dominant genetic model, enrichment of predicted high-impact variants was observed for *ARNTL* (CADD and Maverick OR 234 [95% CI 7·8, 6975]) and *PHKG1* (CADD OR 8·8 [6·5, 12], Maverick OR 10,997 [677, 178,541]) using both prioritization algorithms, though with very wide confidence intervals reflecting sparse counts of qualifying variants (Supplemental Table 15).

## Discussion

In the largest multi-population GWAS meta-analysis of neuropathy to date, we identified seven new loci associated with neuropathy in individuals with type 2 diabetes, including intronic variants in *PHGDH* and *PSPH*, which encode two of the three key enzymes involved in de novo serine biosynthesis. Multiple complementary analyses including GWAS, pathway enrichment, and rare variant burden testing in inherited neuropathy independently converged on the serine biosynthesis pathway as a driver of neuropathy risk, linking diabetic and inherited neuropathies through a shared metabolic vulnerability.

Critically, Mendelian randomization analyses supported a causal relationship of serine levels and neuropathy. Together, these findings establish a genetically informed framework for investigating serine metabolism as a target for therapeutic development in DPN.

Serine occupies a central position in several essential metabolic and biosynthetic pathways, including carbohydrate and lipid metabolism, folate-mediated one-carbon metabolism, and protein synthesis.^34^ Although classified as a non-essential amino acid, serine becomes conditionally essential in tissues with high demand for protein synthesis and lipid homeostasis. Pathogenic variants in *PHGDH, PSAT1* and *PSPH*, which encode the three enzymes of the de novo serine biosynthesis pathway, are known to cause a spectrum of neurodevelopmental and neuroretinal disorders, including progressive neuropathies and macular telangiectasia type 2 (MacTel)^34,35^ (Figure 4). Notably, all three serine biosynthesis genes, were implicated in the present study, either directly through genome-wide significant associations (*PHGDH* and *PSPH*) or indirectly through a trans-eQTL linking the lead *PHGDH* variant to *PSAT1* expression.

**Figure 4.**
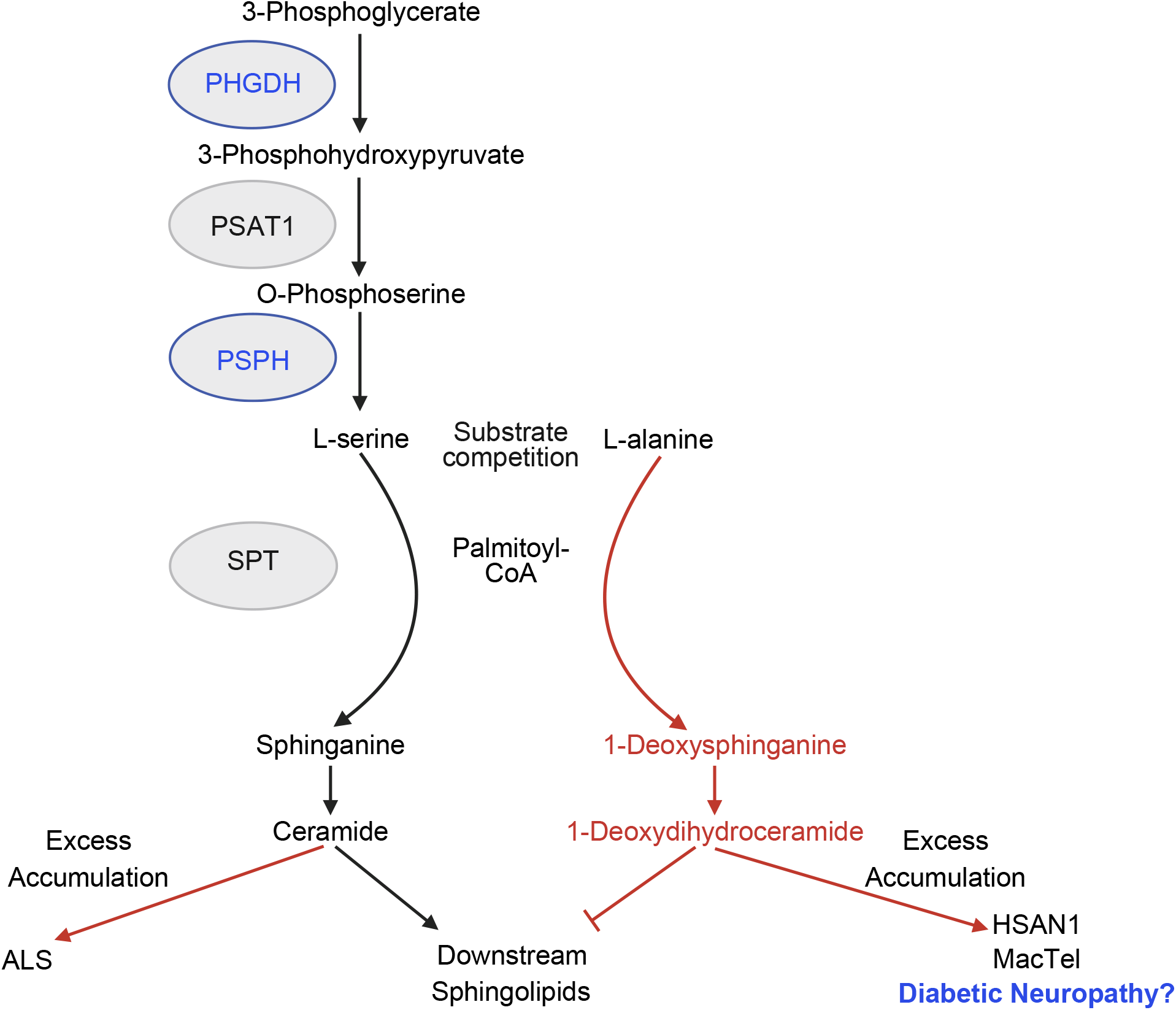
Schematic of the serine and downstream sphingolipid synthetic pathways and associated neuroretinal disorders. The current GWAS findings suggest a potential relationship between the serine biosynthetic pathway and DPN, thereby linking DPN to other serine and sphingolipid metabolism-associated diseases. Abbreviations: MVP, Million Veterans Project; UKB, United Kingdom Biobank; AoU, All of Us; Palmitoyl-CoA, palmitoyl coenzyme-A; SPTLC1, Serine Palmitoyltransferase Long Chain Subunit 1; SPTLC2, Serine Palmitoyltransferase Long chain subunit 2; HSAN1, Hereditary Sensory and Autonomic Neuropathy type 1; ALS, Amyotrophic Lateral Sclerosis; MacTel, Macular Telangiectasia type 2.

Previous GWAS of MacTel similarly identified genetic variants in *PHGDH* and *PSPH*, including variants in strong linkage disequilibrium with those associated with DPN in the current study.^36–38^ Together, these findings suggest that DPN and MacTel may share serine pathway vulnerability.

In addition to impaired serine synthesis, altered serine utilization has been implicated in neurological disorders. Mutations in *serine palmitoyl transferase* (*SPT*), the enzyme that catalyzes serine substrate in the first step of de novo sphingolipid synthesis, can cause hereditary sensory and autonomic neuropathy type 1A (HSAN1A), MacTel, and amyotrophic lateral sclerosis (ALS).^12,35,39^ In HSAN1A and MacTel, disease causing *SPT* variants alter the enzyme’s substrate specificity, shifting utilization from serine to alanine and resulting in the accumulation of neurotoxic 1-deoxysphingolipids.^12,35^ We and others have previously demonstrated reduced circulating plasma serine and increased 1-deoxysphingolipid concentrations in individuals with diabetes and DPN.^8,34^ The identification of common DPN risk loci in *PHGDH* and *PSPH* in this study, provides genetic support for the hypothesis that an accumulation of variants in key serine synthesis genes, together with metabolic factors influencing serine availability, may contribute to neuropathy susceptibility in diabetes.

The serine and sphingolipid metabolic pathways can be targeted therapeutically.^35,40–43^ In pre-clinical studies, dietary serine supplementation, and modulation of SPT activity, has improved neuropathy phenotypes in obese, diabetic mouse models, and ameliorated retinopathy in heterozygous *PHGDH* mice.^42,44^ In humans, serine supplementation effectively reduced circulating 1-deoxysphingolipids in a clinical trial of HSAN1^43^ and is currently being evaluated in a larger cohort of affected individuals (NCT06113055, Clinicaltrials.gov). Reports in individual patients with *PSPH* mutations show improved neuropathy and wound healing after serine supplementation, suggesting serine metabolism may also be a therapeutic target for diabetic foot ulcers.^45^ While targeting the serine pathway in DPN will likely prove more complex than in Mendelian disorders, our findings open an avenue for previously unexplored treatment targets.

Several additional loci identified in his study contain genes with biological relevance to neuropathy. TEAD1, a primary transcription factor in the Hippo signaling pathway, plays a key role in myelin development and regeneration and is required for effective interactions between non-myelinating Schwan cells and nociceptive axons.^46^ *CYP4F2* and *CYP4F11* are of particular interest given the proposed roles of cytochrome P450 enzymes in oxidative stress, inflammation and neural regeneration, processes implicated in the oxidative injury-mediated pathogenesis of DPN.^47,48^ Furthermore, CYP4F2 was recently shown to play a key role in mitigating 1-deoxysphingolipid toxicity via ω-hydroxylation, offering functional support for its potential role in neuropathy.^49^ Several candidate genes within the chromosome 7 locus also warrant further investigation. For example, *CHCHD2* encodes an interspace membrane protein and together with *CHCHD10*, contributes to efficient mitochondrial respiration in motor neurons.^48^ In highlighting these genetic regions, our findings provide a roadmap for prioritizing plausible pathways for mechanistic studies in DPN.

Our study has several important limitations. First, neuropathy case definitions differed across contributing biobanks, and detailed neuropathy phenotyping beyond diagnostic codes was unavailable, limiting our ability to distinguish diabetic neuropathy from hereditary or idiopathic neuropathies, particularly within MVP. Reassuringly, the identified associations remained robust in sensitivity analyses using more stringent neuropathy definitions and after exclusion of individuals with hereditary neuropathies from UKB case cohorts. Second, the MVP was predominantly male, and race/ethnicity representation was uneven across cohorts, although the study was not restricted to individuals of European ancestry.

Differences in phenotype definition, sample size, and population composition may explain the lack of replication of previously reported neuropathy loci. Finally, although GWAS alone cannot identify causal variants, multiple independent analyses converged on the same pathway and candidate genes, strengthening confidence in the biological relevance of *PHGDH, PSPH*, and serine biosynthesis and providing a foundation for future fine-mapping and functional studies.

By integrating large-scale biobank data with an independent inherited neuropathy cohort, we have identified novel genetic contributors to DPN and uncovered convergent genetic evidence implicating the serine biosynthetic pathway in neuropathy susceptibility. The concordance of common and rare variant analyses, pathway enrichment, and Mendelian randomization suggests that diabetic and inherited neuropathies may share a serine pathway vulnerability. Although the underlying mechanisms require further investigation, these results provide a genetically supported and biologically grounded framework for understanding DPN and prioritize serine metabolism as a biologically plausible pathway for therapeutic development. More broadly, this work demonstrates how large-scale human genetics can uncover biologically actionable mechanisms for common neurological diseases that currently lack disease-modifying treatments.

## Supporting information

Supplemental MVP Core Publication Acknowledgement

## Acknowledgements

This work utilized the Alpine high performance computing resource at the University of Colorado Boulder. Alpine is jointly funded by the University of Colorado Boulder, the University of Colorado Anschutz, Colorado State University, and the National Science Foundation (award 2201538). This research is based on data from the Million Veteran Program, Office of Research and Development, Veterans Health Administration, and was supported by awards MVP009/MVP037 I01-BX005831 and MVP051. This publication does not represent the views of the Department of Veteran Affairs or the United States Government. MVP participants went through a counseling process before they enrolled and provided consent to have their electronic health records reviewed. The VA Central Institutional Review Board gave approval for the study protocol in accordance with the principles of the Declaration of Helsinki. Please see Supplemental Acknowledgements for full list of VA MVP personnel. S.R., J.Z., and S.Z. are the guarantors of this work; they had access to data included in the study and take responsibility for the integrity of the data and the accuracy of the data analysis. We would like to acknowledge all of the participants of the biobanks that were used in this study. We would also like to acknowledge Jacob Bockhorst for help with manuscript preparation and Danelle Carter for her contribution of Figure 4. Finally, we would like to thank Marina Kennerson, Barbara Stranger, Leslie Lange and Ethan Lange for their thoughtful feedback.

## Contributors

V.F and S.R were responsible for study conception. V.F., L.v.d.V, J.Z., S.Z., J.R., and S.R., were responsible for study design and project oversight. S.R, A.K., L.v.d.V., A.J., A.W., and S.R., were responsible for data curation analyses. S.R was responsible for visualization. V.F. and S.R. were responsible for writing the original draft and editing the manuscript. All authors contributed to critical review and revision of the manuscript, and all approve of the manuscript submission. V.F., S.R., L.S.P., S.Z., and J.R. contributed to funding support for the study.

## Data Sharing Statement

GWAS summary statistics from MVP will be deposited in the Database of Genotypes and Phenotypes (dbGaP accession number phs001672.v11·p1) prior to publication. Data from UK Biobank are available to researchers with genuine research inquiries with institutional review board oversight and UK Biobank approval (www.ukbiobank.ac.uk/enable-your-research/apply-for-access). The All of Us Research Program data is publicly available to researchers upon application with verification of completion of mandatory research training and regulatory oversight (www.researchallofus.org/register/).

## SUPPLEMENTAL FIGURE LEGENDS

**Supplemental Figure 1**. Schematic of analyses performed in the study. Abbreviations: EUR, European-ancestry; AFR, African-ancestry; HIS, Hispanic; UKB, UK Biobank; GW, genome-wide; BMI, Body mass index; eQTL, expression quantitative trait locus; IV, instrumental variable.

**Supplemental Figure 2**. Regional association plots for seven genome-wide significant loci associated with neuropathy in individuals with diabetes based on GWAS without adjustment for BMI. Coloring of variants based on linkage disequilibrium with the lead SNP at the locus using 1000 Genomes, all ancestry reference panel.

**Supplemental Figure 3**. Miami plot showing results of multi-population genome-wide association meta-analysis of neuropathy in MVP and UK Biobank in models without adjustment for body mass index (BMI) (above the x-axis) and with adjustment for BMI (below the x-axis) in individuals without diabetes **(A)**. Manhattan plot showing results of multi-population genome-wide association meta-analysis of neuropathy in MVP individuals with type 1 diabetes (**B**). Red dotted line indicates the threshold for genome-wide significance (*p* < 5×10^−8^).

**Supplemental Figure 4**. Miami plot showing results of multi-population genome-wide association meta-analysis of neuropathy in MVP and UK Biobank in models without adjustment for body mass index (primary GWAS analysis) (above the x-axis) and sensitivity analysis using a more stringent case definition (below the x-axis) in individuals with diabetes. Red dotted line indicates the threshold for genome-wide significance (*p* < 5×10^−8^).

**Supplemental Figure 5**. Enrichment among tissue-specific differentially expressed gene sets (DEG) across 30 tissue types in GTEx v8 of the neuropathy-associated genes based on physical location of genome-wide significant variants and eQTL associations. Red bars indicate significant enrichment based on *P* <0·05 after multiple testing correction. Separate differentially expressed gene sets evaluated based on up-regulation (top panel), down-regulation (middle panel), and agnostic to direction of differential expression (bottom panel).

**Supplemental Figure 6. (A)** Results of gene set enrichment analysis examining 34,771 gene sets in the Human Molecular Signatures Database. Two gene sets with significant enrichment and their component genes are shown; red dotted line indicates Bonferroni-corrected threshold of significance (*p* < 1·44×10^−6^ based on 34,771 gene sets tested), and the blue dotted line indicates nominal significance (*p* < 0·05). **(B)** Results of custom gene set enrichment analysis for three investigator-defined gene sets comprised of genes with a relationship to neuropathy and serine synthesis (Gene Set A), genes associated with macular telangiectasia and neuropathic conditions (Gene Set B), genes related to glycine synthesis (Gene Set C). Three gene sets with significant enrichment and their component genes are shown; red dotted line indicates Bonferroni-corrected threshold of significance (*p* < 0·01 based on 5 custom gene sets tested), and the blue dotted line indicates nominal significance (*p* < 0·05).

**Supplemental Figure 7**. Bayesian colocalization analysis evaluating overlap of GWAS associations with neuropathy in individuals with diabetes and with plasma serine levels in Lotta et al (*Nature Genetics* 2021; 53(1):54-64) at two loci associated with both traits at genome-wide significance, *PHGDH* **(A)** and *PSPH* **(B)**. Plots show SNP associations with neuropathy (top) and with serine levels (bottom); in each plot, color corresponds to linkage disequilibrium (LD) with the lowest p-value SNP at the locus for each trait, and square-shaped SNPs are members of the 95% credible set of variants based on posterior probability for causality for both neuropathy and serine levels at the locus. Colocalization results for each locus are quantified as the posterior probability in support of five hypotheses: *H*_*0*_ that the data do not support a causal genetic association with either trait; *H*_*1*_ that the data support a causal genetic association for trait 1 (neuropathy) but not trait 2 (serine levels); *H*_*2*_ that the data support a causal genetic association for trait 2 (serine) but not trait 1 (neuropathy); *H*_*3*_ that the data support a causal genetic association for both traits but without a shared causal variant; and *H*_*4*_ that the data support a shared causal genetic variant for both traits. For each SNP analyzed, *coloc* also provides the posterior probability that the variant is the shared causal variant for both traits assuming *H*_*4*_ is true, and the “credible set” of variants whose posterior probabilities sum to >0·95 are shown for each locus.

**Supplemental Figure 8**. Sensitivity analyses of two-sample Mendelian randomization (MR) testing causal association of plasma serine levels with diabetic peripheral neuropathy based on metabolome GWAS in Lotta et al (*Nature Genetics* 2021; 53(1):54-64) and multi-population GWAS meta-analysis of neuropathy in individuals with type 2 diabetes. (**A**) Consistency of effect of serine-associated single nucleotide polymorphisms (SNPs) on neuropathy and lines representing MR effects across five methods. (**B**) Consistency of MR-estimated causal association of plasma serine levels with neuropathy in individuals with type 2 diabetes across five MR estimation methods. Odds ratios shown indicate genetically-estimated neuropathy risk per genetically-estimated standard deviation increase in plasma serine levels. (**C**) Leave-one-out analysis evaluating influence of each genetic instrument (SNP) in the MR analysis. Odds ratios shown indicate genetically-estimated neuropathy risk per genetically-estimated standard deviation increase in plasma serine levels when all five variants are included in the analysis (“All SNPs”) and when the indicated SNP is omitted from the MR analysis.

