## Supplemental Methods, Supplemental Figures 1-8, & Supplemental Tables 1-15 for "Convergent human genetic evidence implicates serine biosynthesis in diabetic peripheral neuropathy"

**Table of Contents**

**Online Supplemental Methods**

1. *Data sources and study participants*
2. *Genomic data information*
3. *GWAS summarization*
4. *Gene-based analysis*
5. *Gene set enrichment analysis*
6. *Rare, high-impact variants in individuals with inherited neuropathy*
7. *Investigating association of serine with neuropathy*

**Online Supplemental Tables**

**Supplemental Table 1.** Mapping Phecodes to ICD-9 and ICD-10 diagnosis codes for neuropathy outcomes used in GWAS in MVP.
**Supplemental Table 2.** Primary care, ICD-9, and ICD-10 diagnosis codes used to define neuropathy outcome in UK Biobank.
**Supplemental Table 3.** Genes in custom gene sets related to serine biosynthesis, neuropathy, glycine biosynthesis, and/or macular telangiectasia.
**Supplemental Table 4.** Independent significant variants nominated by FUMA and COJO at each genome-wide significant locus associated with neuropathy in individuals with type 2 diabetes.
**Supplemental Table 5.** Associations with neuropathy in individuals without diabetes of GWAS-significant variants associated with neuropathy in individuals with type 2 diabetes.
**Supplemental Table 6.** Top genome-wide significant associations with neuropathy in individuals with type 2 diabetes in multi-population meta-analysis without and with adjustment for BMI.
**Supplemental Table 7.** Sensitivity of top genome-wide significant associations with neuropathy in individuals with type 2 diabetes to a stringent case definition.
**Supplemental Table 8.** Replication of genome-wide associated loci in All of Us.
**Supplemental Table 9.** GWAS associations of variants previously reported to be associated with neuropathy or neuropathic pain.
**Supplemental Table 10.** GWAS associations with neuropathy of variants previously reported to be associated with macular telangiectasia.
**Supplemental Table 11.** Genes mapped to genome-wide significant SNPs in individuals with type 2 diabetes based on genomic position and eQTL associations.
**Supplemental Table 12.** Enrichment of genome-wide association signals in gene sets related to neuropathy, serine biosynthesis, and/or macular telangiectasia.
**Supplemental Table 13.** Characteristics of genetic instrumental variables drawn from plasma metabolite GWAS used for Mendelian randomization analyses.
**Supplemental Table 14.** Sensitivity of rare variant burden test associations with inherited neuropathy to variant prioritization stringency.
**Supplemental Table 15.** Association of rare, predicted pathogenic variants with inherited neuropathy case status under a dominant variant filtering and genetic model.

**References**

**Online Supplemental Methods**
*1. Data sources and study participants*

Biobank-scale data for this study came from three cohorts, each of which has been previously described: The design of the Million Veteran Program (MVP) has been previously described.[1,2] Participants were recruited from >60 VA medical centers starting in 2011; those who provided written informed consent were enrolled and provided a blood sample from which DNA was extracted for genotyping. MVP phenotype data is derived from the VA electronic health record (EHR). Ethical and human subjects research oversight for the MVP and this specific project were provided by the VA Central Institutional Review Board. We included MVP enrollees through 2023, stratified into three mutually exclusive cohorts analyzed in parallel: those with type 2 diabetes, based on at least one use of an ICD-9 or ICD-10 diagnosis code for type 2 diabetes and no use of ICD-9 or ICD-10 codes for type 1 diabetes; those with type 1 diabetes, based on at least one use of an ICD-9 or ICD-10 diagnosis code for type 1 diabetes; and those without diabetes, defined as the absence of any ICD-9 or ICD-10 diagnosis codes for diabetes of any type.

The UK Biobank is a research study that has enrolled half a million individuals for genotyping and linkage of genetic data with data from hospital records, as well as primary care data linkage in a subset. Voluntary participants provided informed consent and then participated in a study examination and provided responses to demographic, socioeconomic, and health-related questions.[3] UKB participants were stratified into two cohorts: those with type 2 diabetes, based on participant self-report or any use of a primary care, ICD-9, or ICD-10 diagnosis code for type 2 diabetes; and those without diabetes, defined as participants without self-reported diabetes of any kind and no use of a primary care, ICD-9, or ICD-10 diagnosis code for any form of diabetes in all linked primary care and hospital records.

The All of Us Research Program (AoU) is a multi-center study that links clinical data with genotyping to facilitate genetic discovery and to accelerate medical breakthroughs that lead to individualized prevention, treatment, and clinical care.[4,5] In this study, we examined AoU for replication of the primary GWAS meta-analysis in individuals with type 2 diabetes. AoU participants with diabetes mellitus were identified on the basis of OMOP concept IDs 201254, 201826, 43531011, 4008576, 442793, 443238, 4016045, 43531007, 45757079, and their descendant concepts. Among those with diabetes, type 1 diabetes (T1D) were defined as those with ≥2 T1D codes, ≥50% of diabetes codes specific to T1D, prescribed insulin at any time, and either prescribed glucagon at any time or not prescribed oral glucose-lowering medications. Type 2 diabetes (T2D) patients were defined as having ≥2 T2D-specific codes at least 30 days apart and not being classified as T1D.

*2. Genomic data*

Genotyping, genotyping quality control, and imputation in the MVP have been previously described.[1,2] Genotyping of DNA extracted from blood was performed on a customized Affymetrix Axiom biobank array (MVP 1·0 Genotyping Array). Samples were excluded if duplicated, if they had excess heterozygosity, if there were >2·5% of missing genotype calls, or in the presence of sex-gender discordance. One individual from each pair of related individuals was removed. Variants were excluded on the basis of poor calling (posterior call probability <0·9, call rate <97·5% for common variants [MAF >1%]) or allele frequency discrepancies with 1000 Genomes Project reference data.[6] Genotypes were imputed into the MVP from the TransOmics for Precision Medicine (TOPMed) Program reference panel.[7] After imputation, variants were excluded for the following criteria: ancestry-specific Hardy-Weinberg equilibrium[8] *p*-value $<1x10^{-20}$, imputation quality <0.3, and minor allele frequency (MAF) <0·01. Finally, global and ancestry-specific principal components (PCs) were calculated using flashPCA software to adjust for population stratification.

Genotyping in the UK Biobank has been described elsewhere.[9] Briefly, genotyping was performed on all participants using the UK Biobank Axiom array, including ~850,000 directly genotyped variants with imputation to the UK10K and 1000 Genomes Phase 3 reference panels. Sample quality control included discarding mismatches of genetic sex and self-reported sex, sex chromosome aneuploidy, and outliers for heterozygosity or missing calls. Variants were excluded for MAF <0·001 and for imputation info score <0·3. Genotyping in AoU has been described previously.[5] Briefly, we used short-read whole-genome sequencing (WGS) data from the AoU controlled tier dataset V8. Short-read WGS were generated from blood-derived DNA using PCR-free Illumina KAPA HyperPrep libraries sequenced on Illumina NovaSeq 6000 instruments, with initial QC performed with the Illumina DRAGEN pipeline.

*3. GWAS summarization*

GWAS results were summarized using FUMA (http://fuma.ctglab.nl/), a platform that annotates, prioritizes, and visualizes GWAS results.[10] Independent, genome-wide significant SNPs were defined as those with $p<5\text{x}10^{-8}$ and with LD $r^2<0{\cdot}6$ with each other. SNPs with $p<0{\cdot}05$ were grouped into a genomic locus if they were linked at $r^2\geq 0{\cdot}6$ or were physically close (distance <250kb). Lead SNPs were defined within each locus if they were independent ($r^2<0.1$) and genome-wide significant ($p<5\text{x}10^{-8}$). We used LD score regression (LDSC) to estimate SNP-based heritability from the GWAS summary statistics,[11] and we performed conditional joint analysis (COJO) to evaluate for multiple, independent signals at the genome-wide significant loci.[12]

*4. Gene-based analysis*

We used Multimarker Analysis of GenoMic Annotation (MAGMA) to perform gene-based association tests with neuropathy case status.[13] We first mapped all SNPs included in the GWAS to genes based on physical location. MAGMA then uses a multiple linear principal components model to test for associations of genes (based on summary statistics of SNP-trait associations) with case status. The MAGMA gene-based test accounts for LD between SNPs mapped to a gene and yields a gene-based association p-value that is identifiable even in the presence of collinear SNPs.[13] We performed the gene-based analysis for each population-specific GWAS and for results of the multi-population meta-analysis.

*5. Gene set enrichment analysis*

We also used MAGMA to perform gene set enrichment analysis (GSEA), a statistical tool to assess whether related genes (aggregated into gene sets) harbor statistical evidence of association with the trait of interest in the absence of independent, genome-wide SNP associations in those genes.[13] In MAGMA, GSEA compares the association statistics from gene-based tests for gene sets to a null distribution for a gene set of similar size. We performed GSEA using a catalog of >34,000 gene sets available from Molecular Signatures Database (gsea-msigdb.org/gsea/msigdb/)[14,15] and using five custom gene sets: Set A. Genes with a relationship to neuropathy and serine biosynthesis; Set B. Genes associated with macular telangiectasia (a hereditary condition with neuropathic features) and other neuropathic conditions; Set C. Genes related to glycine biosynthesis (based on connecting glycine and serine synthesis); Set D. Genes with a relationship to serine biosynthesis but not to neuropathy; Set E. Genes associated with macular telangiectasia but not known to be related to serine biosynthesis (Supplemental Table 3). Genes were identified through review of the literature[16–31] and of the serine biosynthesis pathway in the Kyoto Encyclopedia of Genes and Genomes Pathway Database (KEGG)[32], and use of tools such as GeneCards[33] and Uniprot[34] to confirm relationships of a gene with serine biosynthesis.

*6. Rare, high-impact variants in individuals with inherited neuropathy*

The *GENESIS* database comprises more than 20,000 rare-disease genomic datasets contributed by academic consortia and individual investigators.[35] As of November 2025, 2,519 individuals in this cohort were diagnosed with Charcot-Marie-Tooth disease (CMT) or related hereditary motor and sensory neuropathies; among them, 2,099 were classified as unsolved, defined as having no molecular diagnosis after expert clinical genetics review. Population controls were obtained from the AoU cohort after exclusion of individuals carrying ICD-9 or ICD-10 codes indicating any form of neuropathy, resulting in 245,394 control individuals.

For gene-burden analyses, we interrogated 11 genes highlighted in the GWAS results: *PHGDH, PSPH, SUMF2, CCT6A, PHKG1, CHCHD2, TEAD1, ARNTL, CYP4F2, CYP4F11,* and *LARGE1*. To enrich for rare, plausibly pathogenic variants, we implemented two filtering strategies based on the CADD PHRED score or Maverick score.[36,37] Both included standard depth and quality metrics (depth [DP] > 12, genotype quality [GQ] > 50, Quality [QUAL] > 175), and restricted functional consequences on MANE Select transcripts to splice acceptor, splice donor, stop-gain, stop-loss, frameshift, start-loss, in-frame deletion, in-frame insertion, or missense variants. Predicted functional impact was based on the CADD PHRED score[33] or Maverick score[37], which also accounts for the modeled mode of inheritance.

The primary analysis examined whether carrying a predicted high-impact variant was associated with hereditary neuropathy status similar to an additive model employed in GWAS. For this analysis, we used variants with gnomAD v2 allele frequency (AF) $< 0{\cdot}01$ and either CADD PHRED $\geq 10$ or a Maverick recessive score $\geq 0{\cdot}5$. As a secondary analysis, we examined predicted high-impact variants that may impact neuropathy risk in a dominant fashion. For this analysis, we used variants with gnomAD v2 AF $< 0{\cdot}001$ and either a CADD PHRED score $\geq 20$ or a Maverick dominant score of $\geq 0{\cdot}5$.

For each gene and model (additive or dominant, high impact based on CADD or Maverick), variant sets were constructed independently in the Genesis and AoU cohorts using identical criteria. Carrier counts were then used to compute odds ratios (ORs) for association with case status. Standard errors were estimated using the Delta method, and p-values were adjusted for multiple testing using the Benjamini-Hochberg correction.

*7. Investigating association of serine with neuropathy*

We performed three analyses to evaluate the relationship of serine and neuropathy using genetic tools. First, we estimated genetic correlation of plasma serine levels and DPN by applying LD Score Regression[38] to publicly available summary statistics from a GWAS of plasma metabolites[39] and the European ancestry GWAS summary statistics for DPN in MVP. Second, for two loci, *PHGDH* (chr 1) and *PSPH* (chr 7), with genome-wide significant associations with both DPN and plasma serine levels, we applied Bayesian colocalization analysis in the R package *coloc* to estimate the posterior probability in support of a hypothesis that both traits share a causal genetic variant.[40] Again, we used publicly available summary statistics from a recent GWAS of plasma serine levels in this analysis.[39] Third, we performed two-sample Mendelian randomization (MR) to test for evidence supporting a causal association of serine levels with DPN using genetic instruments for plasma serine levels drawn from six independent genetic studies in diverse populations.[39,41–45] Using the *TwoSampleMR* R package, the primary analysis was based on the invariance variance weighted method, and robustness of MR results was examined using tests for genetic instrument heterogeneity, pleiotropy (MR-Egger), and sensitivity to omission of variants in the genetic instrument.[46,47]

**Online Supplemental Tables**

**Supplemental Table 1.** Mapping Phecodes to ICD- 9 and ICD-10 diagnosis codes for neuropathy outcomes used in GWAS in MVP.

| Phecode | ICD-9 codes | ICD-10 codes |
|---|---|---|
| **337·1** | 337·0<br>337·00<br>337·09<br>337·1 | G99·0<br>G90·0<br>G90·09 |
| **356** | 356<br>356·0<br>356·1<br>356·2<br>356·3<br>356·4<br>356·8<br>356·9 | G60·8<br>G60·9<br>G60·2<br>G60·0<br>G60·3<br>G60·1<br>G60 |
| **250·6** | 357·2 | E10·42<br>E08·42<br>E09·42 |
| **250·14** | 250·61<br>250·63 | E10·40<br>E10·4<br>E10·41<br>E10·42<br>E10·43<br>E10·44<br>E10·49 |
| **250·24** | 250·60<br>250·62 | E11·4<br>E11·40<br>E11·41<br>E11·42<br>E11·43<br>E11·44<br>E11·49 |

**Supplemental Table 2.** Primary care, ICD-9, and ICD-10 diagnosis codes used to define neuropathy outcome in UK Biobank.

| Vocabulary | DPN (Type 2 Diabetes) | Neuropathy (Non-diabetes) | Hereditary Neuropathy |
|---|---|---|---|
| **ICD10** | E104, E114, E134, E144, G578, G579, G587, G589, G590, G600, G603, G608, G609, G629, G632, G633, G638, G900, G990, M146 | G578, G579, G587, G589, G600, G603, G608, G609, G629, G633, G638, G900, G990, M146 | G602 |
| **ICD9** | 2505, 3370, 3371, 3561, 3562, 3564, 3568, 3569, 3572 | 3370, 3371, 3561, 3562, 3564, 3568, 3569 | |
| **Read Codes** | 1M8.., C106., C1060, C1061, C106y, C106z, C1092, C10FB, C10FH, F170., F170z, F171., F1711, F171z, F35z., F35z0, F36.., F364., F366., F367., F36y., F36yz, F36z., F372., F3720, F3721, F3722, F37y1, F3y0., Fyu6D, Fyu7., Fyu70, Fyu72, Fyu7C, FyuAC, M2711, M2712, M2717, N0301, N035., N2423, SD722, X00A8, X00Ag, X00Ah, X00Aj, X00Al, X00Am, X00CF, X00CG, X00CH, X00CI, X50Bi, X50Bj, X702P, X75sy, Xa0lK, Xa0YH, Xa1G9, Xa3fB, Xa8Hz, XaA1H, XaB3q, XaBtQ, Xac0x, XaEno, XaEnp, XaEnq, XaFmM, XaFn9, XaPmX, XaXbW, XC0eJ, XE10H, XE12I, XE15i, XE15k, XE18L, XE2sp, Y0c4c, Y0c4d, Y2667, Y2668, YA345 | F170., F170z, F171., F171z, F35z., F36.., F364., F366., F367., F36y., F36yz, F36z., F37y1, Fyu7., Fyu70, Fyu72, Fyu7C, M2717, N035., N2423, SD722, X00A8, X00CF, X00CG, X00CH, X00CI, X50Bi, X50Bj, X702P, X75sy, Xa0YH, Xa1G9, Xa3fB, Xa8Hz, XaA1H, XaB3q, XaBtQ, XC0eJ, XE15i, XE18L, XE2sp, Y0c4c, Y0c4d, Y2667, Y2668, YA345 | F360z, F361., F3610, F361z, F362., F368., F3680, F3681, X00A9, X00AA, X00AM, X00AO |
| **UKB self-reported condition codes** | 1255, 1468 | 1255 | |

**Supplemental Table 3.** Genes in custom gene sets related to serine biosynthesis, neuropathy, glycine biosynthesis, and/or macular telangiectasia.

| **Gene Set A** | | **Gene Set B** | | **Gene Set C** | | **Gene Set D** | | **Gene Set E** | |
|---|---|---|---|---|---|---|---|---|---|
| **Gene** | **Locus** | **Gene** | **Locus** | **Gene** | **Locus** | **Gene** | **Locus** | **Gene** | **Locus** |
| *AKT1* | 14q32·33 | *MFN2* | 1p36·22 | *ALDH1L1* | 3q21·3 | *AOC2* | 17q21·31 | *CERS4* | 19p13·2 |
| *ALDH7A1* | 5q23·2 | *CHCHD2* | 7p11·2 | *MTHFD1* | 14q23·3 | *DAO* | 12q24·11 | *CPS1* | 2q34 |
| *CBS* | 21q22·3 | *DCTN1* | 2p13·1 | *MTHFD2* | 2p13·1 | *KDM6A* | Xp11·3 | *EOMES* | 3p24·1 |
| *CCT6A* | 7p11·2 | *SCN11A* | 3p22·2 | *SHMT1* | 17p11·2 | *KDSR* | 18q21·33 | *FBN3* | 19p13·2 |
| *CPS1* | 2q34 | *LARS2* | 3p21·31 | *SHMT2* | 12q13·3 | *PGK1* | Xq21·1 | *GBAS* | 7p11·2 |
| *GAMT* | 19p13·3 | *SLC25A46* | 5q22·1 | *MTR* | 1q43 | *SLC7A5* | 16q24·2 | *HMGCS2* | 1p12 |
| *GRIN1* | 9q34·3 | *SLC4A7* | 3p24·1 | *GLDC* | 9q24·1 | *SPTSSA* | 14q13·1 | *JMJD1C* | 10q21·3 |
| *GRIN2B* | 12p13·1 | | | *GART* | 21q22·11 | *SPTSSB* | 3q26·1 | *LIMD1* | 3p21·31 |
| *ORMDL1* | 2q32·2 | | | *ATIC* | 2q35 | *THNSL1* | 10p12·1 | *NRBF2* | 10q21·3 |
| *ORMDL1* | 2q32·2 | | | *MTHFD1L* | 6q25·1 | *TMEM161B* | 5q14·3 | *REEP3* | 10q21·3 |
| *ORMDL2* | 12q13·2 | | | *MTMFT* | 15q22·31 | | | *SLC6A20* | 3p21·31 |
| *ORMDL3* | 17q21·1 | | | *ALDH1L2* | 12q23·3 | | | *SUMF2* | 7p11·2 |
| *PGAM1* | 10q24·1 | | | *TYMS* | 18p11·32 | | | *TMEM161B* | 5q14·3 |
| *PHGDH* | 1p12 | | | | | | | *TTC39B* | 9p22·3 |
| *PSAT1* | 9q21·2 | | | | | | | *ZNF713* | 7p11·2 |
| *PSPH* | 7p11·2 | | | | | | | | |
| *SDS* | 12q24·13 | | | | | | | | |
| *SLC1A4* | 2p14 | | | | | | | | |
| *SLC1A5* | 19q13·32 | | | | | | | | |
| *SPTLC1* | 9q22·31 | | | | | | | | |
| *SPTLC2* | 14q24·3 | | | | | | | | |
| *SRR* | 17p13·3 | | | | | | | | |

Set A: Genes with a relationship to neuropathy and serine biosynthesis
Set B: Genes associated with macular telangiectasia and neuropathic conditions
Set C: Genes related to glycine biosynthesis
Set D: Genes with a relationship to serine biosynthesis but not to neuropathy.
Set E: Genes associated with macular telangiectasia but not known to be related to serine biosynthesis.

**Supplemental Table 4.** Independent significant variants nominated by FUMA and COJO at each genome-wide significant locus associated with neuropathy in individuals with type 2 diabetes.

| **Nearest gene** | **Chr** | **Position** | **rsID** | **Effect allele** | **EAF** | **OR** | **p-value** | **Effect direction (NHW/NHB/HIS/AS/ UKB)** | **COJO nominated independent variant** |
|---|---|---|---|---|---|---|---|---|---|
| *PHGDH* | 1 | 120254545 | rs562038 | C | 0·71 | 0·95 | 4·1e-12 | -/-/-/+/- | Y |
| | | 120267597 | rs3838425 | G | 0·45 | 1·04 | 9·3e-12 | +/+/+/-/na | N |
| | | 120254506 | rs561931 | A | 0·40 | 1·04 | 1·1e-10 | +/+/+/-/+ | N |
| | | 120224469 | rs866321 | T | 0·36 | 1·04 | 2·5e-8 | +/+/+/-/na | N |
| *COBLL1* | 2 | 165501849 | rs3923113 | A | 0·58 | 1·04 | 1·0e-8 | +/+/+/-/+ | Y |
| *PSPH* | 7 | 56110766 | rs2016724 | C | 0·52 | 1·03 | 8·8e-9 | +/+/+/+/+ | N |
| | | 56109770 | rs2002651 | C | 0·52 | 1·03 | 8·9e-9 | +/+/+/+/+ | Y |
| *TEAD1* | 11 | 12866146 | rs2099744 | A | 0·63 | 0·96 | 1·2e-9 | -/-/-/-/- | Y |
| *FTO* | 16 | 53803223 | rs62048402 | A | 0·38 | 1·05 | 2·2e-14 | +/+/+/+/+ | Y |
| | | 53822169 | rs113935429 | A | 0·40 | 1·05 | 2·2e-13 | +/+/+/+/na | N |
| | | 53824226 | rs62033406 | A | 0·62 | 0·96 | 4·3e-13 | -/-/-/-/- | N |
| | | 53799977 | rs9930333 | T | 0·56 | 0·96 | 4·0e-10 | -/-/-/-/- | N |
| | | 53807764 | rs17817288 | A | 0·53 | 0·97 | 1·4e-9 | -/-/-/-/- | N |
| | | 53829963 | rs201399553 | T | 0·62 | 0·96 | 3·7e-9 | -/-/-/-/na | N |
| | | 53807005 | rs10718688 | A | 0·54 | 1·04 | 7·9e-9 | +/+/+/+/na | N |
| | | 53797908 | rs7206790 | C | 0·51 | 0·97 | 1·4e-8 | -/-/-/-/na | N |
| | | 53804340 | rs1861866 | T | 0·56 | 1·03 | 2·1e-8 | +/+/+/+/+ | N |
| *CYP4F11* | 19 | 16041150 | rs11670222 | C | 0·59 | 1·04 | 2·4e-8 | +/+/+/-/na | N |
| | | 16033701 | rs11451979 | T | 0·59 | 1·03 | 3·0e-8 | +/+/+/-/na | Y |
| *LARGE1* | 22 | 34312130 | rs13058212 | A | 0·002 | 11·2 | 2·4e-8 | na/na/na/na/+ | Y |

**Supplemental Table 5.** Associations with neuropathy in individuals without diabetes of GWAS-significant variants associated with neuropathy in individuals with type 2 diabetes.

| Nearest gene | Chr | Position | rsID | Effect allele | EAF | OR | p-value | Effect direction (NHW/NHB/HIS/AS/UKB) |
|---|---|---|---|---|---|---|---|---|
| *PHGDH* | 1 | 120254545 | rs562038 | C | 0·71 | 0·97 | 2·4e-4 | ---+- |
| *COBLL1* | 2 | 165501849 | rs3923113 | A | 0·59 | 1·01 | 4·3e-1 | +++-+ |
| *PSPH* | 7 | 56110766 | rs2016724 | C | 0·51 | 1·01 | 6·5e-2 | ++--- |
| *TEAD1* | 11 | 12866146 | rs2099744 | A | 0·66 | 0·99 | 1·4e-1 | -+-+- |
| *FTO* | 16 | 53803223 | rs62048402 | A | 0·37 | 1·01 | 2·5e-1 | +-+-+ |
| *CYP4F11* | 19 | 16041150 | rs11670222 | C | 0·57 | 1·01 | 2·3e-1 | ++--? |
| *LARGE1* | 22 | 34312130 | rs13058212 | A | 0·002 | 0·99 | 9·7e-1 | ????- |

Abbreviations: Chr, chromosome; EAF, effect allele frequency; OR, odds ratio; NHW, non-Hispanic White; NHB, non-Hispanic Black; HIS, Hispanic; AS, Asian; UKB, UK Biobank; na, variant data not available

**Supplemental Table 6.** Top genome-wide significant associations with neuropathy in individuals with type 2 diabetes in multi-population meta-analysis without and with adjustment for BMI.

| | | | | | | Without BMI adjustment | | | With BMI adjustment | | |
|---|---|---|---|---|---|---|---|---|---|---|---|
| **Nearest gene** | **Chr** | **Position** | **rsID** | **Effect allele** | **EAF** | **OR** | **p-value** | **Effect direction (NHW/NHB/HIS/AS/UKB)** | **OR** | **p-value** | **Effect direction (NHW/NHB/HIS/AS/UKB)** |
| *PHGDH* | 1 | 120254545 | rs562038 | C | 0·71 | 0·95 | 4·1e-12 | -/-/-/+/- | 0·95 | 5·4e-12 | -/+/-/+/- |
| *COBLL1* | 2 | 165501849 | rs3923113 | A | 0·58 | 1·04 | 1·0e-8 | +/+/+/-/+ | 1·04 | 3·9e-11 | +/+/+/-/+ |
| *PSPH* | 7 | 56110766 | rs2016724 | C | 0·52 | 1·03 | 8·8e-9 | +/+/+/+/+ | 1·04 | 2·3e-9 | +/+/+/+/+ |
| *TEAD1* | 11 | 12866146 | rs2099744 | A | 0·63 | 0·96 | 1·2e-9 | -/-/-/-/- | 0·96 | 1·7e-9 | -/-/-/+/- |
| *FTO* | 16 | 53803223 | rs62048402 | A | 0·38 | 1·05 | 2·2e-14 | +/+/+/+/+ | 1·03 | 1·3e-7 | +/+/+/+/+ |
| *CYP4F11* | 19 | 16041150 | rs11670222 | C | 0·59 | 1·04 | 2·4e-8 | +/+/+/-/na | 1·04 | 8·2e-9 | +/+/+/-/na |
| *LARGE1* | 22 | 34312130 | rs13058212 | A | 0·002 | 11·2 | 2·4e-8 | na/na/na/na/+ | 12·1 | 1·4e-8 | na/na/na/na/+ |
| *HLA-DQA1* | 6 | 32591588 | rs9271608 | A | 0·83 | 0·96 | 2·7e-7 | -/-/-/-/- | 0·95 | 3·6e-9 | -/-/-/-/- |
| *TCF7L2* | 10 | 114758349 | rs7903146 | T | 0·32 | 1·02 | 3·7e-4 | +/+/+/-/+ | 1·04 | 9·0e-11 | +/+/+/-/+ |

**Supplemental Table 7.** Sensitivity of top genome-wide significant associations with neuropathy in individuals with type 2 diabetes to a stringent case definition.

| | | | | | | Primary analysis | | | Sensitivity analysis | | |
|---|---|---|---|---|---|---|---|---|---|---|---|
| **Nearest gene** | **Chr** | **Position** | **rsID** | **Effect allele** | **EAF** | **OR** | **p-value** | **Effect direction (NHW/NHB/HIS/ AS/UKB)** | **OR** | **p-value** | **Effect direction (NHW/NHB/HIS/ AS/UKB)** |
| *PHGDH* | 1 | 120254545 | rs562038 | C | 0·71 | 0·95 | 4·1e-12 | -/-/-/+/- | 0·94 | 5·2e-15 | -/+/-/+/- |
| *COBLL1* | 2 | 165501849 | rs3923113 | A | 0·58 | 1·04 | 1·0e-8 | +/+/+/-/+ | 1·04 | 2·7e-8 | +/+/+/-/+ |
| *PSPH* | 7 | 56110766 | rs2016724 | C | 0·52 | 1·03 | 8·8e-9 | +/+/+/+/+ | 1·04 | 2·0e-8 | +/+/+/-/+ |
| *TEAD1* | 11 | 12866146 | rs2099744 | A | 0·63 | 0·96 | 1·2e-9 | -/-/-/-/- | 0·96 | 3·4e-9 | -/-/-/+/- |
| *FTO* | 16 | 53803223 | rs62048402 | A | 0·38 | 1·05 | 2·2e-14 | +/+/+/+/+ | 1·06 | 3·7e-17 | +/+/+/+/- |
| *CYP4F11* | 19 | 16041150 | rs11670222 | C | 0·59 | 1·04 | 2·4e-8 | +/+/+/-/na | 1·04 | 1·1e-9 | +/+/+/-/na |
| *LARGE1* | 22 | 34312130 | rs13058212 | A | 0·002 | 11·2 | 2·4e-8 | na/na/na/na/+ | 11·5 | 1·2e-5 | na/na/na/na/+ |
| *HLA-DQA1* | 6 | 32591588 | rs9271608 | A | 0·83 | 0·96 | 2·7e-7 | -/-/-/-/- | 0·96 | 9·2e-8 | -/-/-/-/- |
| *TCF7L2* | 10 | 114758349 | rs7903146 | T | 0·32 | 1·02 | 3·7e-4 | +/+/+/-/+ | 1·04 | 1·7e-8 | +/+/+/+/+ |

**Supplemental Table 8.** Replication of genome-wide associated loci in All of Us.

| | | | | | EUR | | | AFR | | | AMR | | |
|---|---|---|---|---|---|---|---|---|---|---|---|---|---|
| **Nearest gene** | **Chr** | **Position** | **rsID** | **Effect allele** | **EAF** | **OR** | **p-value** | **EAF** | **OR** | **p-value** | **EAF** | **OR** | **p-value** |
| *PHGDH* | 1 | 120254545 | rs562038 | C | 0·68 | 0·92 | 5·0e-4 | 0·89 | 1·01 | 7·5e-1 | 0·72 | 0·95 | 1·9e-1 |
| *COBLL1* | 2 | 165501849 | rs3923113 | A | 0·65 | 1·03 | 1·8e-1 | 0·32 | 1·02 | 6·0e-1 | 0·76 | 1·03 | 3·8e-1 |
| *PSPH* | 7 | 56110766 | rs2016724 | C | 0·49 | 0·97 | 1·2e-1 | 0·64 | 0·99 | 8·3e-1 | 0·46 | 0·98 | 4·5e-1 |
| *TEAD1* | 11 | 12866146 | rs2099744 | A | 0·72 | 0·94 | 9·3e-3 | 0·39 | 0·98 | 4·9e-1 | 0·75 | 0·93 | 4·9e-2 |
| *FTO* | 16 | 53803223 | rs62048402 | A | 0·44 | 1·10 | 2·6e-6 | 0·10 | 1·11 | 2·8e-2 | 0·25 | 1·00 | 9·6e-1 |
| *CYP4F11* | 19 | 16041150 | rs11670222 | C | 0·53 | 1·05 | 2·3e-2 | 0·86 | 1·06 | 1·1e-1 | 0·63 | 0·99 | 7·0e-1 |
| *LARGE1* | 22 | 34312130 | rs13058212 | A | 0·003 | 1·00 | 9·8e-1 | 4·9e-4 | 0·93 | 9·1e-1 | 4·6e-4 | 1·16 | 8·4e-1 |

Abbreviations: Chr, chromosome; EAF, effect allele frequency; OR, odds ratio; EUR, European ancestry; AFR, African ancestry; AMR, Amerindian ancestry.

**Supplemental Table 9.** GWAS associations of variants previously reported to be associated with neuropathy or neuropathic pain.

| | | | | | | Non-diabetic individuals | | | | Type 2 diabetic individuals | | | | |
|---|---|---|---|---|---|---|---|---|---|---|---|---|---|---|
| **Trait** | **Nearest gene** | **Chr** | **Position** | **rsID** | **Effect allele** | **EAF** | **OR** | **p-value** | **Effect direction (NHW/NHB/HIS/ AS/UKB)** | **EAF** | **OR** | **p-value** | **Effect direction (NHW/NHB/HIS/ AS/UKB)** | **Ref** |
| Idiopathic poly-neuropathy | *B4GALNT3* | 12 | 588944 | rs7294354 | T | 0.49 | 1.01 | 1.4e-1 | +/+/+/-/? | 0.50 | 1.00 | 8.2e-1 | +/-/+/-/? | 48 |
| | *NR5A2* | 1 | 199854673 | rs147738081 | T | 0.03 | 1.07 | 1.2e-2 | +/?/+/?/? | 0.03 | 1.02 | 2.9e-1 | +/?/+/?/? | 48 |
| Diabetic neuropathy | *SCN2A* | 2 | 167629849 | rs13417783 | T | 0.14 | 1.01 | 5.8e-1 | +/+/-/-/? | 0.14 | 0.99 | 4.1e-1 | -/-/+/-/? | 49 |
| Neuropathic pain | *SLC25A3* | 12 | 98585582 | rs369920026 | A | 0.006 | 1.03 | 8.6e-1 | ?/?/?/?/+ | 0.006 | 0.92 | 6.8e-1 | ?/?/?/?/- | 50 |
| | *CAB39L* | 13 | 49905672 | rs7992766 | A | 0.74 | 1.01 | 3.8e-1 | +/+/+/-/? | 0.73 | 1.01 | 1.1e-1 | +/+/+/-/? | 50 |
| Neuropathic pain | *KCNT2* | 1 | 196273664 | rs114159097 | C | 0.03 | 1.00 | 8.9e-1 | -/+/-/-/- | 0.04 | 1.00 | 9.5e-1 | +/-/-/+/- | 51 |
| | *ANK2* | 4 | 113813347 | rs72669682 | A | 0.02 | 0.99 | 6.4e-1 | -/?/?/?/- | 0.02 | 1.00 | 8.8e-1 | +/?/?/?/+ | 51 |
| Diabetic neuropathic pain | *GFRA2* | 8 | 21711431 | rs17428041 | T | 0.75 | 1.01 | 3.4e-1 | +/+/-/-/+ | 0.76 | 1.00 | 5.6e-1 | -/-/+/+/+ | 52 |
| Taxane-induced peripheral neuropathy | *GNGT1* | 7 | 93349015 | rs1858826 | T | 0.91 | 1.00 | 9.1e-1 | +/-/-/?/? | 0.91 | 1.01 | 3.7e-1 | +/-/+/?/? | 53 |
| | *NXN* | 17 | 701122 | rs910920 | A | 0.27 | 1.02 | 7.5e-2 | +/+/+/+/? | 0.27 | 1.00 | 5.4e-1 | -/+/+/+/? | 53 |
| | *MIR5684* | 4 | 165370126 | rs1857798 | T | 0.65 | 1.00 | 8.1e-1 | +/-/-/+/- | 0.66 | 1.00 | 4.6e-1 | -/+/-/-/? | 53 |
| | *FGD2* | 6 | 36979583 | rs12202642 | T | 0.05 | 0.99 | 6.9e-1 | -/+/-/-/? | 0.95 | 1.00 | 9.7e-1 | +/?/-/-/? | 53 |
| Diabetic foot ulcer | *MAPK14, SLC26A8* | 6 | 35993906 | rs3761980 | A | 0.87 | 0.99 | 1.9e-1 | -/+/-/-/- | 0.85 | 1.01 | 3.3e-1 | +/-/-/+/+ | 54 |
| | *MAPK14* | 6 | 35998388 | rs80028505 | T | 0.12 | 1.01 | 3.2e-1 | +/-/-/+/+ | 0.12 | 0.99 | 1.2e-1 | -/-/+/-/- | 54 |
| Diabetic neuropathy | *GYPA* | 4 | 145030546 | rs1132787 | T | 0.27 | 1.00 | 9.1e-1 | -/+/-/-/+ | 0.26 | 1.00 | 9.6e-1 | -/+/-/+/+ | 55 |
| | *LOC105371557* | 17 | 15733545 | rs522521 | A | 0.67 | 1.00 | 7.8e-1 | -/-/-/+/+ | 0.65 | 1.00 | 7.9e-1 | +/-/-/+/- | 55 |

**Supplemental Table 10.** GWAS associations with neuropathy of variants previously reported to be associated with macular telangectasia.

| | | | | | Non-diabetic individuals | | | | Type 2 diabetic individuals | | | | |
|---|---|---|---|---|---|---|---|---|---|---|---|---|---|
| **Nearest gene** | **Chr** | **Position** | **rsID** | **Effect allele** | **EAF** | **OR** | **p-value** | **Effect direction (NHW/NHB/HIS/ AS/UKB)** | **EAF** | **OR** | **p-value** | **Effect direction (NHW/NHB/HIS/ AS/UKB)** | **Ref** |
| *PHGDH* | 1 | 120257576 | rs477992 | A | 0·32 | 1·02 | 2·5e-4 | +/+/+/-/+ | 0·32 | 1·04 | 1·9e-10 | +/-/+/-/+ | 14 |
| | | 120265444 | rs532303 | A | 0·31 | 1·03 | 8·7e-4 | +/+/+/-/+ | 0·31 | 1·04 | 2·2e-9 | +/-/+/-/+ | 20 |
| | | 120278072 | rs146953046 | T | 0·99 | 0·78 | 3·2e-2 | ?/?/?/?/- | 0·99 | 0·83 | 2·1e-1 | ?/?/?/?/- | 20 |
| *SLC1A4* | 2 | 65220910 | rs2160387 | T | 0·53 | 0·99 | 2·7e-1 | -/-/+/+/+ | 0·51 | 1·00 | 7·2e-1 | -/-/+/+/+ | 20 |
| *CPS1* | 2 | 211543055 | rs715 | T | 0·73 | 1·02 | 2·3e-2 | +/-/+/+/? | 0·73 | 1·02 | 8·6e-3 | +/-/-/-/? | 14 |
| | | 211540507 | rs1047891 | A | 0·31 | 0·99 | 6·6e-2 | -/+/-/-/+ | 0·31 | 0·98 | 2·2e-3 | -/-/+/+/- | 20 |
| *SLC4A7; EOMES* | 3 | 27706298 | rs9820465 | T | 0·74 | 0·99 | 1·5e-1 | -/-/-/-/+ | 0·71 | 1·00 | 9·8e-1 | -/+/+/+/- | 20 |
| *SLC6A20* | 3 | 45814094 | rs17279437 | A | 0·09 | 0·99 | 6·5e-1 | -/+/-/-/+ | 0·09 | 0·99 | 2·4e-1 | -/-/+/-/- | 20 |
| *TMEM161B* | 5 | 87786004 | rs73171800 | T | 0·86 | 0·99 | 3·2e-1 | -/-/+/+/- | 0·85 | 1·00 | 9·7e-1 | -/+/+/+/+ | 14 |
| | | 87847586 | rs17421627 | T | 0·93 | 1·00 | 8·3e-1 | -/+/+/+/- | 0·93 | 0·99 | 4·9e-1 | -/-/+/+/+ | 20 |
| *PSPH* | 7 | 56099352 | rs6955423 | A | 0·72 | 0·98 | 2·4e-3 | -/+/+/+/+ | 0·71 | 0·97 | 8·1e-6 | -/-/-/-/- | 20 |
| *TTC39B* | 9 | 15302613 | rs677622 | A | 0·13 | 1·00 | 9·99e-1 | +/-/+/+/- | 0·13 | 1·01 | 1·0e-1 | +/+/+/+/- | 20 |
| *NRBF2* | 10 | 65363166 | rs10995566 | T | 0·31 | 1·01 | 2·6e-1 | +/-/-/-/- | 0·30 | 1·00 | 5·8e-1 | +/+/+/-/+ | 20 |
| *CERS4; FBN3* | 19 | 8235251 | rs139412173 | A | 0·95 | 1·03 | 1·4e-1 | +/-/+/?/- | 0·95 | 1·00 | 9·9e-1 | -/+/-/?/+ | 20 |

**Supplemental Table 11.** Genes mapped to genome-wide significant SNPs in individuals with type 2 diabetes based on genomic position and eQTL associations.

| Gene Symbol | Chr | Entrez ID | Mapped based on position | Mapped based on eQTL | Minimum p-value of eQTL association | eQTL Direction | GWAS Locus |
|---|---|---|---|---|---|---|---|
| WARS2 | 1 | 10352 | - | + | 3·63E-11 | + | *PHGDH* |
| ZNF697 | 1 | 90874 | - | + | 1·30E-19 | + | *PHGDH* |
| PHGDH | 1 | 26227 | + | + | 7·05E-96 | - | *PHGDH* |
| HMGCS2 | 1 | 3158 | - | + | 6·59E-05 | - | *PHGDH* |
| NOTCH2 | 1 | 4853 | - | + | NA | + | *PHGDH* |
| EPHB1 | 3 | 2047 | - | + | 1·57E-06 | - | *PHGDH* |
| PSAT1 | 9 | 29968 | - | + | 1·03E-16 | + | *PHGDH* |
| TMPRSS13 | 11 | 84000 | - | + | 4·65E-08 | - | *PHGDH* |
| SLC7A1 | 13 | 6541 | - | + | 7·16E-09 | + | *PHGDH* |
| SMOC1 | 14 | 64093 | - | + | 6·51E-06 | - | *PHGDH* |
| AARS | 16 | 16 | - | + | 3·27E-07 | + | *PHGDH* |
| EVI2A | 17 | 2123 | - | + | 7·55E-06 | - | *PHGDH* |
| FIGN | 2 | 55137 | - | + | 5·54E-05 | + | *COBLL1-GRB14* |
| GRB14 | 2 | 2888 | - | + | 6·41E-15 | + | *COBLL1-GRB14* |
| COBLL1 | 2 | 22837 | + | + | 1·11E-10 | - | *COBLL1-GRB14* |
| SLC38A11 | 2 | 151258 | - | + | 3·32E-15 | - | *COBLL1-GRB14* |
| SCN2A | 2 | 6326 | - | + | 2·28E-12 | + | *COBLL1-GRB14* |
| VOPP1 | 7 | 81552 | - | + | 2·01E-06 | - | *PSPH* |
| MRPS17 | 7 | 51373 | - | + | 1·34E-17 | - | *PSPH* |
| ZNF713 | 7 | 349075 | - | + | 3·72E-25 | - | *PSPH* |
| GBAS | 7 | 2631 | - | + | 2·01E-28 | + | *PSPH* |
| PSPH | 7 | 5723 | + | + | 5·54E-06 | + | *PSPH* |
| CCT6A | 7 | 908 | + | + | 1·62E-139 | + | *PSPH* |
| SUMF2 | 7 | 25870 | + | + | 1·86E-109 | - | *PSPH* |
| PHKG1 | 7 | 5260 | + | + | 3·86E-19 | - | *PSPH* |
| CHCHD2 | 7 | 51142 | + | + | 3·27e-310 | + | *PSPH* |
| NUPR1L | 7 | 389493 | + | + | 4·76E-22 | + | *PSPH* |
| TEAD1 | 11 | 7003 | + | + | 7·94E-08 | + | *TEAD1* |
| TUBB4B | 9 | 10383 | - | + | 4·21E-06 | + | *FTO* |
| RBL2 | 16 | 5934 | - | + | 1·43E-07 | - | *FTO* |
| FTO | 16 | 79068 | + | + | 8·20E-13 | + | *FTO* |
| IRX3 | 16 | 79191 | - | + | 1·05E-06 | + | *FTO* |
| ZNF480 | 19 | 147657 | - | + | 2·74E-06 | + | *FTO* |
| ILVBL | 19 | 10994 | - | + | 9·65E-06 | + | *CYP4F11* |
| CYP4F22 | 19 | 126410 | - | + | 3·25E-06 | - | *CYP4F11* |
| CYP4F12 | 19 | 66002 | - | + | 1·72E-05 | - | *CYP4F11* |
| CYP4F2 | 19 | 8529 | + | + | 1·25E-23 | + | *CYP4F11* |
| CYP4F11 | 19 | 57834 | + | + | 2·18E-33 | + | *CYP4F11* |
| RAB8A | 19 | 4218 | - | + | 3·73E-07 | + | *CYP4F11* |
| MED26 | 19 | 9441 | - | + | 5·47E-05 | + | *CYP4F11* |
| LARGE | 22 | 9215 | + | - | NA | NA | *LARGE1* |

**Supplemental Table 12.** Enrichment of genome-wide association signals in gene sets related to neuropathy, serine biosynthesis, and/or macular telangiectasia.

| Gene set | Type 2 diabetes |
|---|---|
| A | 4·6e-7* |
| B | 1·6e-4* |
| C | 8·5e-3* |
| D | 0·34 |
| E | 3·4e-2 |

Set A: Genes with a relationship to neuropathy and serine biosynthesis
Set B: Genes associated with macular telangiectasia and neuropathic conditions
Set C: Genes related to glycine biosynthesis
Set D: Genes with a relationship to serine biosynthesis but not to neuropathy.
Set E: Genes associated with macular telangiectasia but not known to be related to serine biosynthesis.
* Significant at a Bonferroni-corrected threshold of 1·0e-2 (0.05/5 gene sets)

**Supplemental Table 13.** Characteristics of genetic instrumental variables drawn from plasma metabolite GWAS used for Mendelian randomization analyses.

| Lead author | Population | Sample size | Variants | Gene region | F-statistic | Heterogeneity p-value | MR Egger intercept (p-value) |
|---|---|---|---|---|---|---|---|
| Chen[41] | European (Canada) | 8,271 | rs1047891<br>rs561931<br>rs816411 | *CPS1 (chr2)*<br>*PHGDH (chr1)*<br>*CHCHD2 (chr7)* | 133·2 | 0·011 | -0·063 (0·43) |
| Lotta[39] | European (UK) | 9,324 | rs477992<br>rs4947534<br>rs28601761<br>rs715<br>rs1260326 | *PHGDH (chr1)*<br>*PSPH (chr7)*<br>*TRIB1AL (chr8)*<br>*CPS1 (chr2)*<br>*GCKR (chr2)* | 112·6 | 0·13 | -0·004 (0·76) |
| Tahir[42] | Black (US) | 2466 | rs477992<br>rs34016595<br>rs1047891 | *PHGDH (chr1)*<br>*ZNF713 (chr7)*<br>*CPS1 (chr2)* | 52·6 | 0·29 | 0·050 (0·37) |
| Feofanova[43] | Multi-population (US) | 11,812 | rs10923893<br>rs1047891<br>rs62265227<br>rs11238389 | *PHGDH (chr1)*<br>*CPS1 (chr2)*<br>*ALDH1L1 (chr3)*<br>*PSPH (chr7)* | 132·8 | 0·45 | 0·013 (0·47) |
| Surendran[44] | European (UK) | 14,296 | rs561931<br>rs1260326<br>rs1047891<br>rs4470984<br>rs28601761<br>rs2465219 | *PHGDH (chr1)*<br>*GCKR (chr2)*<br>*CPS1 (chr2)*<br>*NIPSNAP2 (chr7)*<br>*TRIB1AL (chr8)*<br>*SLC38A4-AS1 (chr12)* | 129·1 | 0·008 | 0·017 (0·23) |
| Imaizumi[45] | Japanese | 1,338 | rs12613336<br>rs13244654 | *CPS1 (chr2)*<br>*SUMF2 (chr7)* | 47·3 | 0·06 | NA[a] |

[a] MR Egger requires a minimum of 3 variants in genetic instrument to estimate intercept.

**Supplemental Table 14.** Sensitivity of rare variant burden test associations with inherited neuropathy to variant prioritization stringency. Odds ratios for association of gene burden of variants with inherited neuropathy case status under the specified variant prioritization filter (CADD score ranging from 5 to 15; Maverick score ranging from 0·1 to 0·5).

| | | **CADD Prioritization, Carrier[a]** | | | **Maverick Prioritization, Carrier[a]** | | |
|---|---|---|---|---|---|---|---|
| **Gene** | **Chr** | **CADD 5** | **CADD 10** | **CADD 15** | **Mav 0.10** | **Mav 0.25** | **Mav 0.5** |
| *PHGDH* | 1 | 12·39 | 12·67 | 12·21 | 14·68 | 13·20 | 9·90 |
| *PSPH* | 7 | 3·37 | 8·54 | 9·07 | 16·12 | 18·47 | 24·98 |
| *PHKG1* | 7 | 4·82 | 4·83 | 4·67 | 13·22 | 13·88 | 21·19 |
| *CCT6A* | 7 | 0·39 | 0·39 | 0·39 | 0·94 | 0·24 | 0·96 |
| *SUMF2* | 7 | 0·41 | 0·37 | 0·33 | 1·19 | 1·41 | 1·30 |
| *CHCHD2* | 7 | 0·12 | 0·12 | 0·12 | 0·56 | 1·89 | 9·74 |
| *TEAD1* | 11 | 0·12 | 0·12 | 0·12 | 0·56 | 1·27 | 14·61 |
| *CYP4F2* | 19 | 0·28 | 0·28 | 0·32 | 0·69 | 0·56 | 0·75 |
| *CYP4F11* | 19 | 0·34 | 0·29 | 0·36 | 0·67 | 0·76 | 0·78 |
| *LARGE1* | 22 | 0·0070 | 0·0070 | 0·0088 | 0·023 | 0·032 | 0·042 |
| | | **CADD Prioritization, Dominant[b]** | | | **Maverick Prioritization, Dominant[b]** | | |
| **Gene** | **Chr** | **CADD 15** | **CADD 20** | **CADD 25** | **Mav 0.1** | **Mav 0.25** | **Mav 0.5** |
| *PHGDH* | 1 | 0·34 | 0·36 | 0·50 | NA[c] | NA[c] | NA[c] |
| *PSPH* | 7 | 0·032 | 0·039 | 0·073 | NA[c] | NA[c] | NA[c] |
| *PHKG1* | 7 | 7·26 | 8·80 | 0·22 | 37·64 | 95·51 | NA[c] |
| *CCT6A* | 7 | 0·24 | 0·38 | 0·20 | 1·00 | 0·97 | 1·21 |
| *SUMF2* | 7 | 0·037 | 0·038 | 0·064 | 2·66 | NA[c] | NA[c] |
| *CHCHD2* | 7 | 0·34 | 0·19 | 0·35 | 0·73 | 1·01 | 0·34 |
| *TEAD1* | 11 | 0·25 | 0·12 | 0·071 | 0·95 | 1·09 | 0·65 |
| *ARNTL* | 11 | 233·93 | 233·93 | NA[c] | 233·93 | 233·93 | NA[c] |
| *CYP4F2* | 19 | 0·014 | 0·020 | 0·14 | NA[c] | NA[c] | NA[c] |
| *CYP4F11* | 19 | 0·017 | 0·026 | 0·11 | 29·23 | NA[c] | NA[c] |
| *LARGE1* | 22 | 0·021 | 0·025 | 0·070 | 0·65 | NA[c] | NA[c] |

[a] Carrier allele frequency < 0·01

[b] Dominant allele frequency < 0·001

[c] No carriers of prioritized variants among cases and/or controls, so association not quantifiable.

**Supplemental Table 15.** Association of rare, predicted pathogenic variants with inherited neuropathy case status under a dominant variant filtering and genetic model.

| | | CADD Prioritization | | | Maverick Prioritization[b] | | |
|---|---|---|---|---|---|---|---|
| **Gene** | **Chr** | **OR** | **95% CI** | **p-value** | **OR** | **95% CI** | **p-value** |
| *PHGDH* | 1 | 0·36 | 0·02, 5·72 | 0·65 | NA[c] | | |
| *PSPH* | 7 | 0·039 | 0·002, 0·62 | 1·1e-5 | NA[c] | | |
| *PHKG1* | 7 | 8·79 | 6·45, 11·98 | 9·5e-25 | NA[c] | | |
| *CCT6A* | 7 | 0·38 | 0·14, 1·01 | 5·1e-2 | 1·21 | 0·17, 8·65 | 0·57 |
| *SUMF2* | 7 | 0·038 | 0·002, 0·61 | 8·8e-6 | NA[c] | | |
| *CHCHD2* | 7 | 0·20 | 0·028, 1·42 | 7·8e-2 | 0·34 | 0·021, 5·52 | 0·41 |
| *TEAD1* | 11 | 0·12 | 0·017, 0·88 | 1·0e-2 | 0·65 | 0·091, 4·66 | 1 |
| *ARNTL* | 11 | 234 | 7·8, 6975 | 1·2e-2 | NA[c] | | |
| *CYP4F2* | 19 | 0·020 | 0·001, 0·32 | 2·4e-10 | NA[c] | | |
| *CYP4F11* | 19 | 0·026 | 0·002, 0·41 | 2·6e-8 | NA[c] | | |
| *LARGE1* | 22 | 0·025 | 0·002, 0·40 | 2·4e-8 | NA[c] | | |

[a] CADD criteria: gnomAD v2 allele frequency < 0.001 and CADD PHRED score ≥ 20[36]
[b] Maverick criteria: gnomAD v2 allele frequency < 0.001 and Maverick dominant score ≥ 0·25[37]
[c] No carriers of prioritized variants among cases and/or controls, so association not quantifiable.

#### SUPPLEMENTAL MATERIAL REFERENCES

.

#### Supplemental Figure 1.

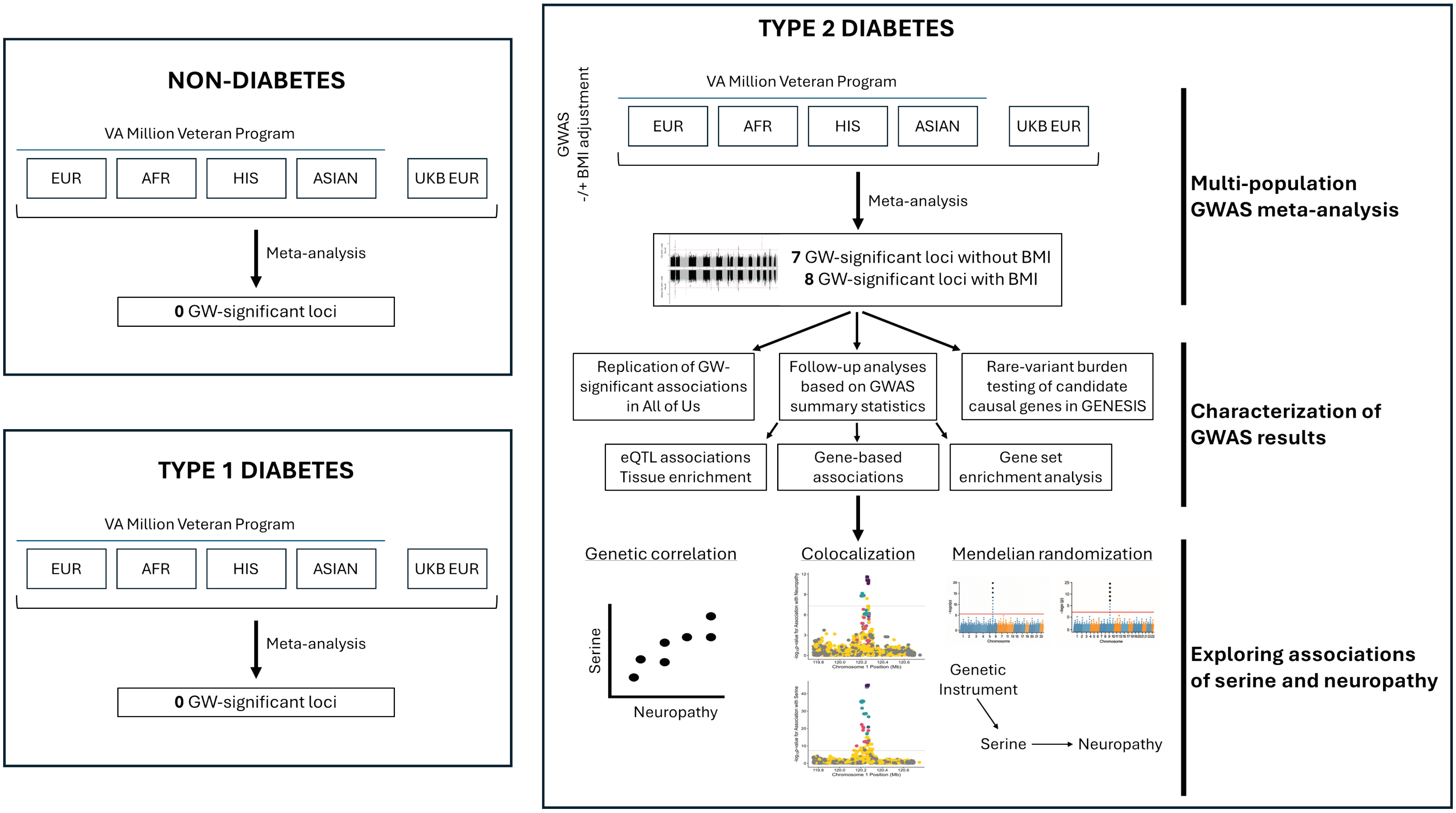


**Supplemental Figure 1.** Schematic of analyses performed in the study. Abbreviations: EUR, European-ancestry; AFR, African-ancestry; HIS, Hispanic; UKB, UK Biobank; GW, genome-wide; BMI, Body mass index; eQTL, expression quantitative trait locus.

### Supplemental Figure 2.

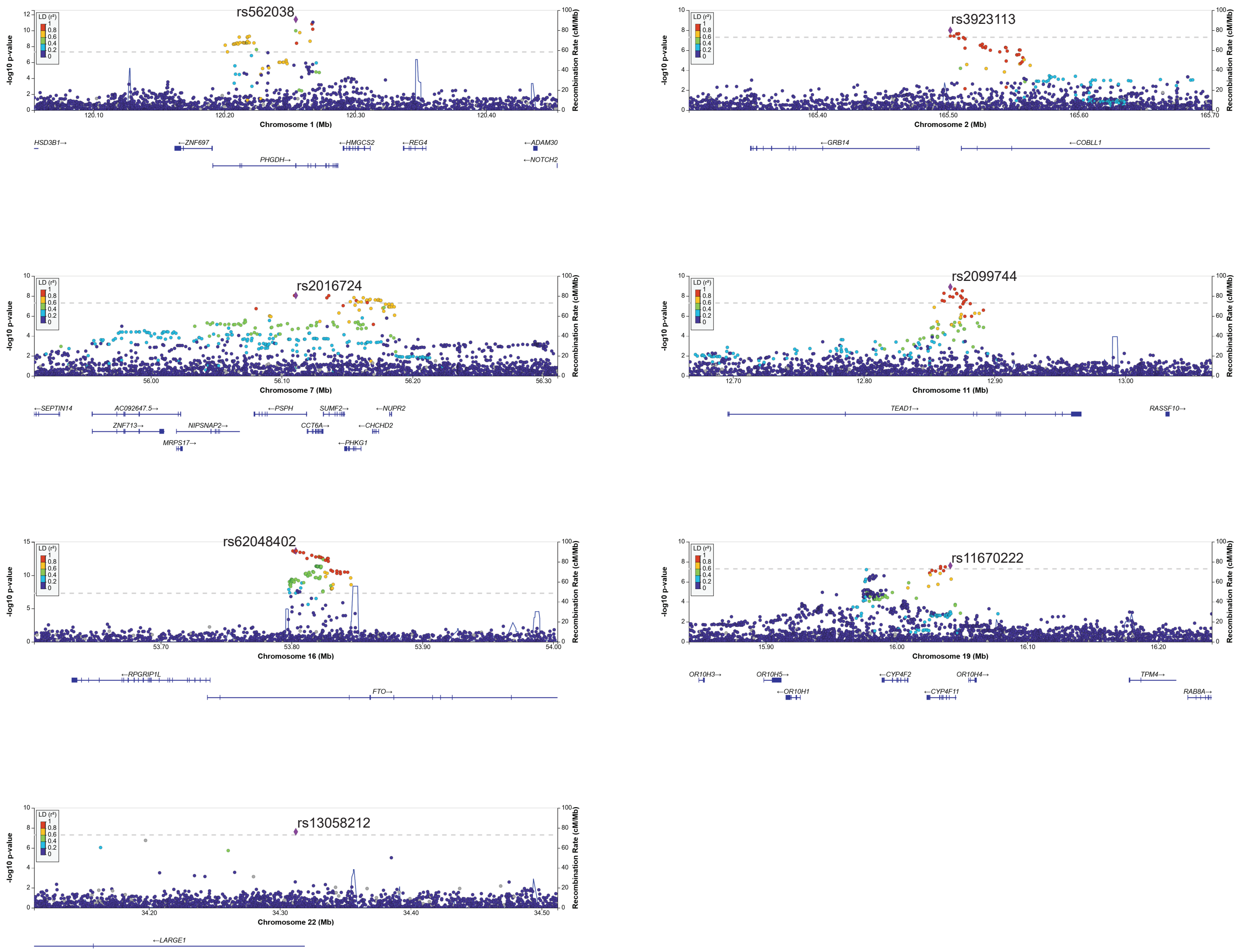


**Supplemental Figure 2.** Regional association plots for seven genome-wide significant loci associated with neuropathy in individuals with diabetes based on GWAS without adjustment for BMI. Coloring of variants based on linkage disequilibrium with the lead SNP at the locus using 1000 Genomes, all ancestry reference panel.

#### Supplemental Figure 3.

**A.**

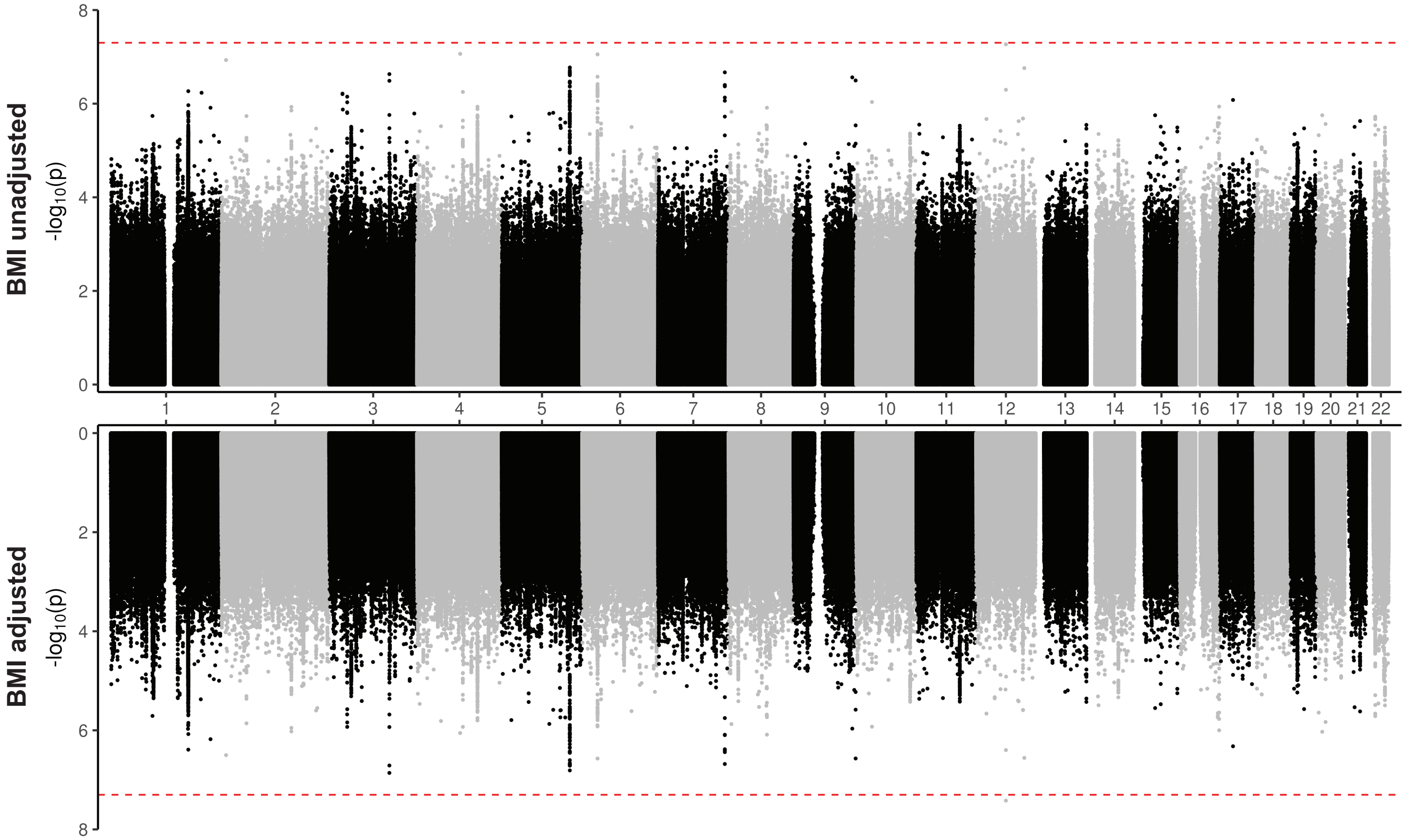


**B.**

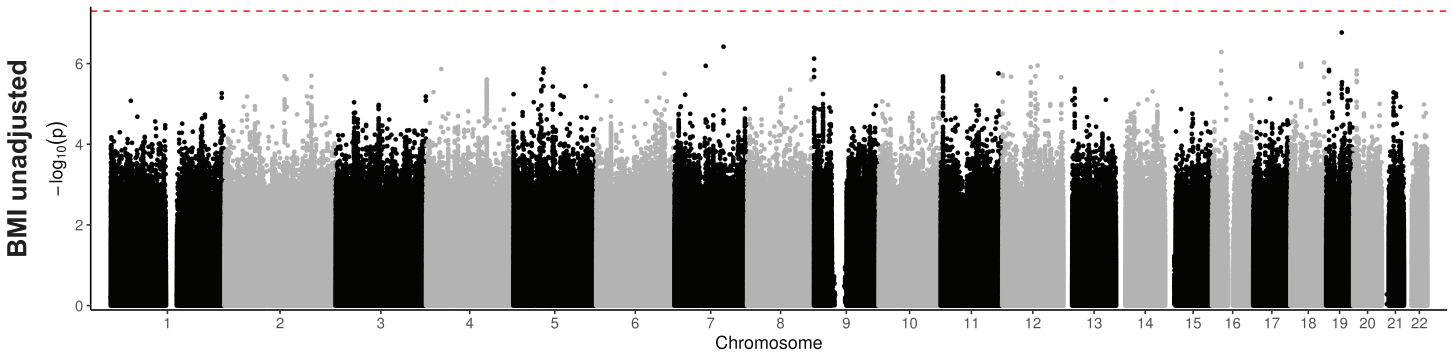


**Supplemental Figure 3.** Miami plot showing results of multi-population genome-wide association meta-analysis of neuropathy in MVP and UK Biobank in models without adjustment for body mass index (BMI) (above the x-axis) and with adjustment for BMI (below the x-axis) in individuals without diabetes **(A)**. Manhattan plot showing results of multi-population genome-wide association meta-analysis of neuropathy in MVP individuals with type 1 diabetes (**B**). Red dotted line indicates the threshold for genome-wide significance ($p < 5x10^{-8}$).

#### Supplemental Figure 4.

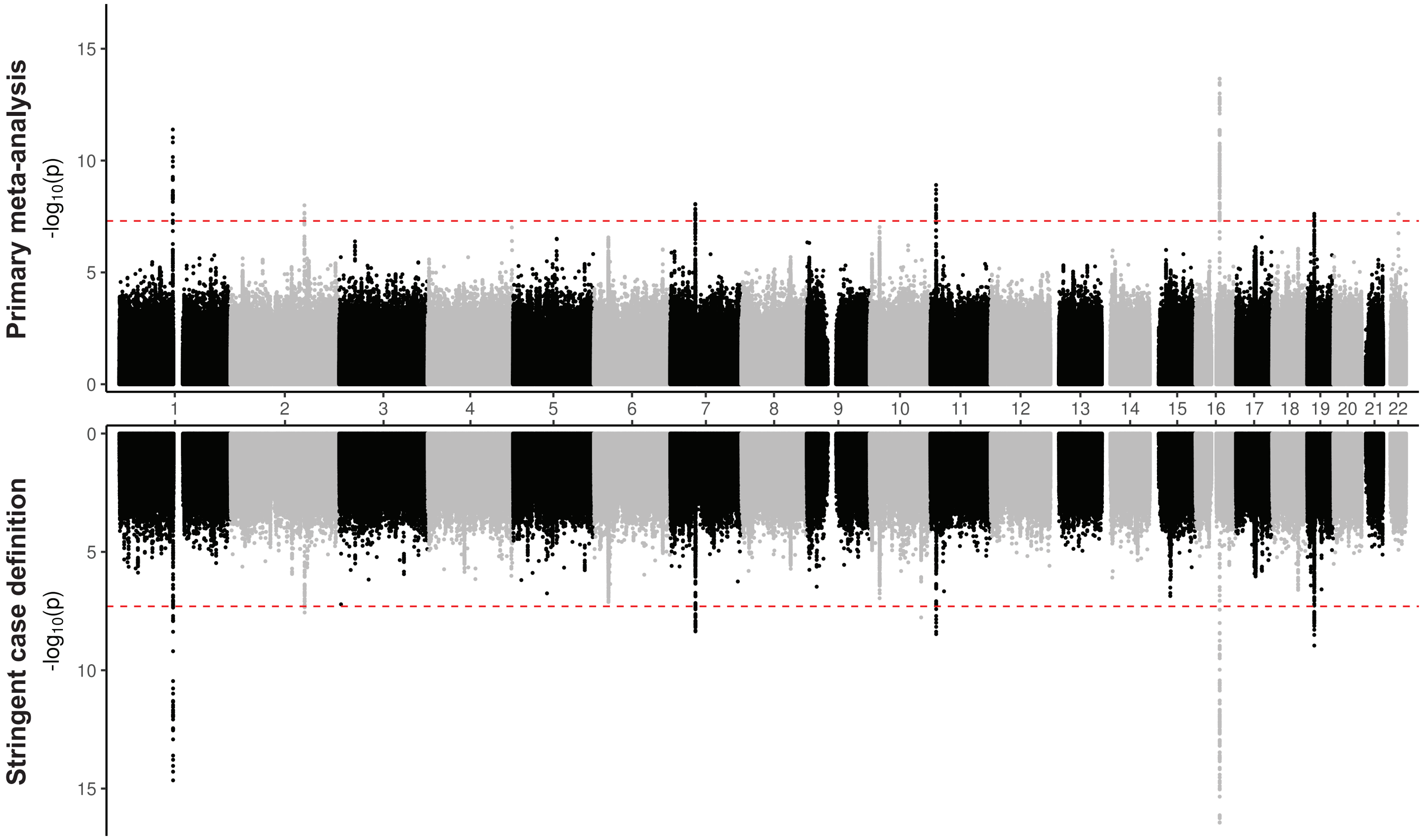


**Supplemental Figure 4.** Miami plot showing results of multi-population genome-wide association meta-analysis of neuropathy in MVP and UK Biobank in models without adjustment for body mass index (primary GWAS analysis) (above the x-axis) and sensitivity analysis using a more stringent case definition (below the x-axis) in individuals with diabetes. Red dotted line indicates the threshold for genome-wide significance ($p < 5x10^{-8}$).

#### Supplemental Figure 5.

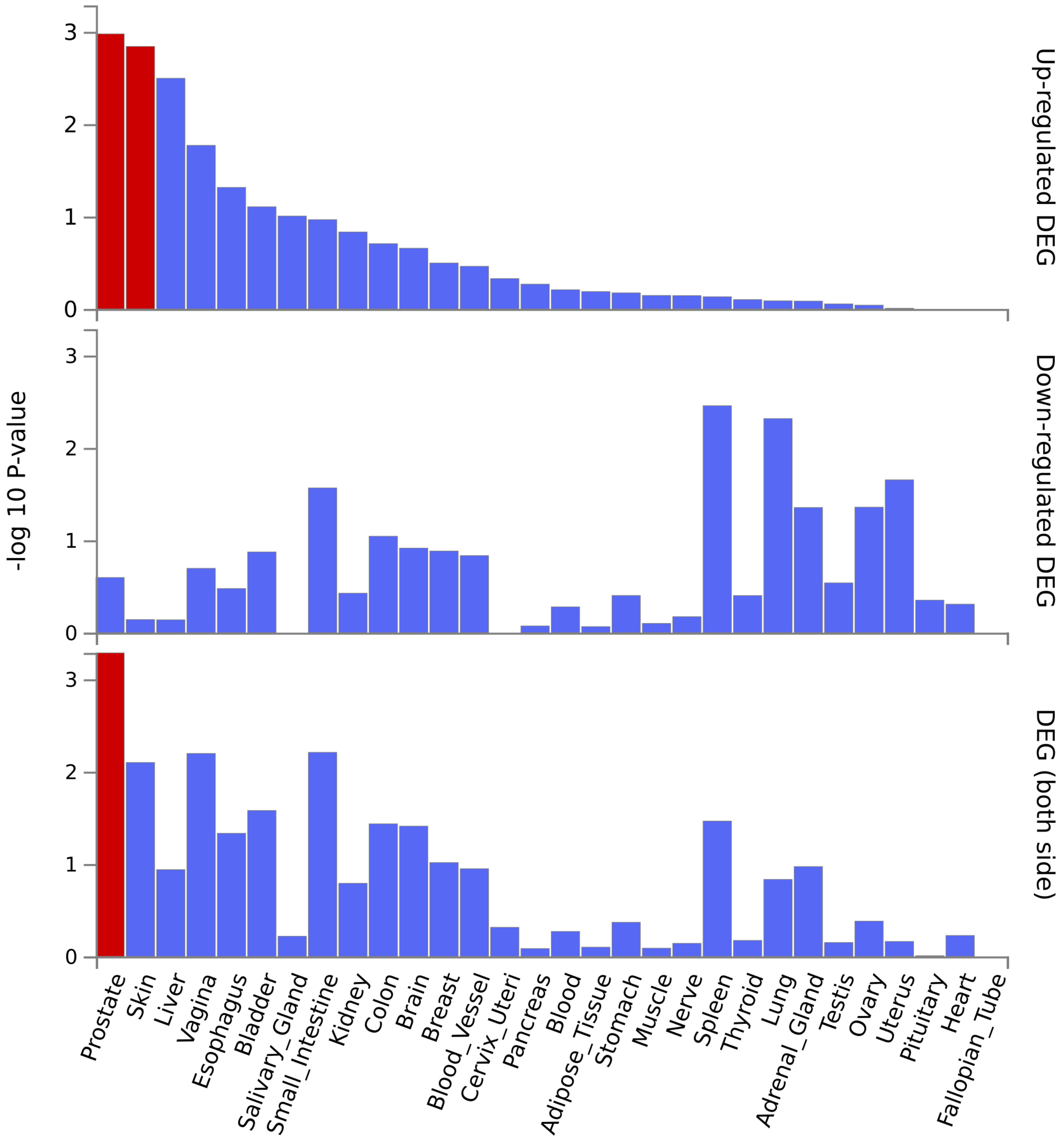


**Supplemental Figure 5.** Enrichment among tissue-specific differentially expressed gene sets (DEG) across 30 tissue types in GTEx v8 of the neuropathy-associated genes based on physical location of genome-wide significant variants and eQTL associations. Red bars indicate significant enrichment based on *P* <0.05 after multiple testing correction. Separate differentially expressed gene sets evaluated based on up-regulation (top panel), down-regulation (middle panel), and agnostic to direction of differential expression (bottom panel).

#### Supplemental Figure 6.

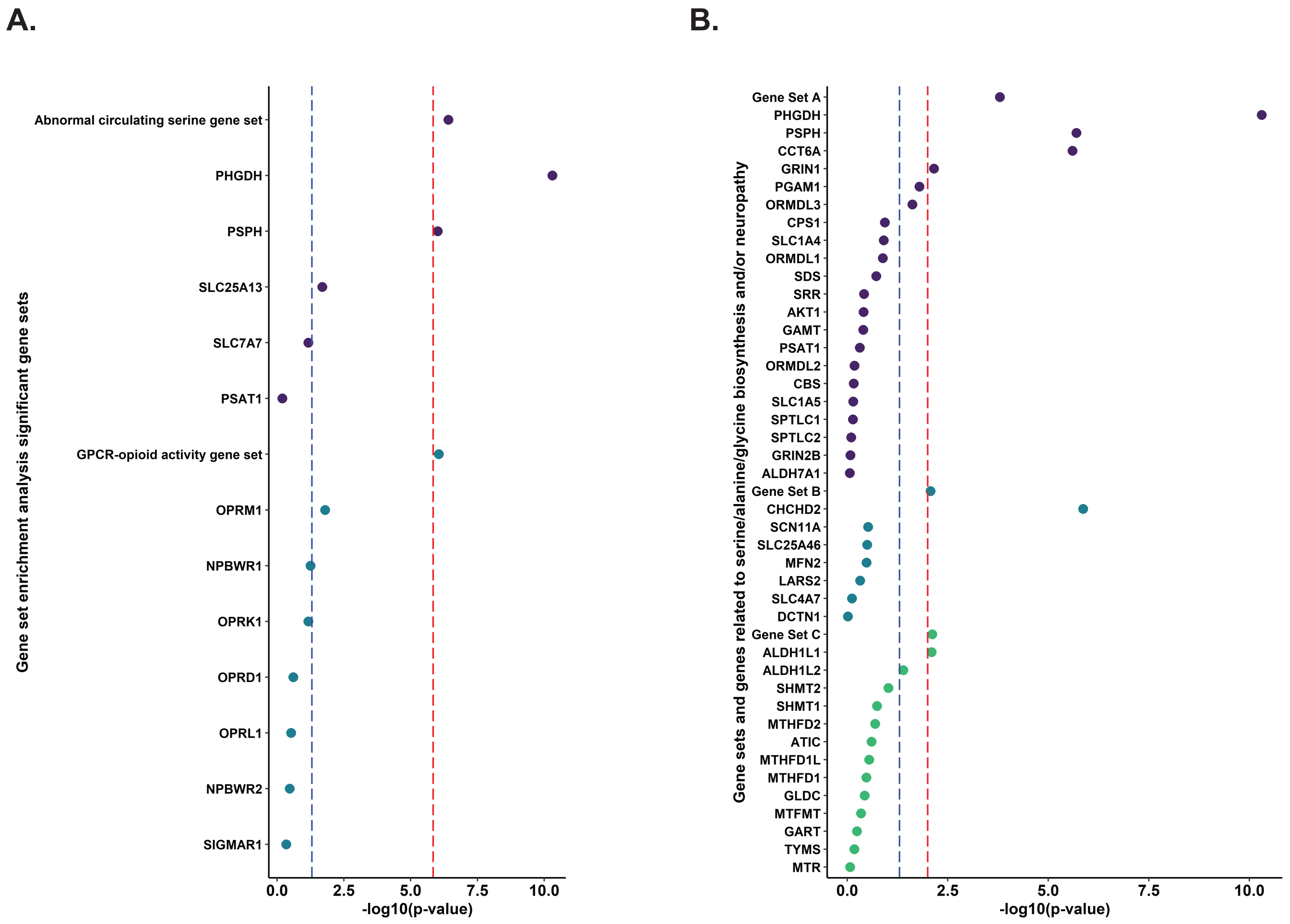


**Supplemental Figure 6. (A)** Results of gene set enrichment analysis examining 34,771 gene sets in the Human Molecular Signatures Database. Two gene sets with significant enrichment and their component genes are shown; red dotted line indicates Bonferroni-corrected threshold of significance ($p < 1.44 \times 10^{-6}$ based on 34,771 gene sets tested), and the blue dotted line indicates nominal significance ($p < 0.05$). **(B)** Results of custom gene set enrichment analysis for three investigator-defined gene sets comprised of genes with a relationship to neuropathy and serine synthesis (Gene Set A), genes associated with macular telangiectasia and neuropathic conditions (Gene Set B), genes related to glycine synthesis (Gene Set C). Three gene sets with significant enrichment and their component genes are shown; red dotted line indicates Bonferroni-corrected threshold of significance ($p < 0.01$ based on 5 custom gene sets tested), and the blue dotted line indicates nominal significance ($p < 0.05$).

#### Supplemental Figure 7.

**A.** Colocalization at *PHGDH* locus

**B.** Colocalization at *PSPH* locus

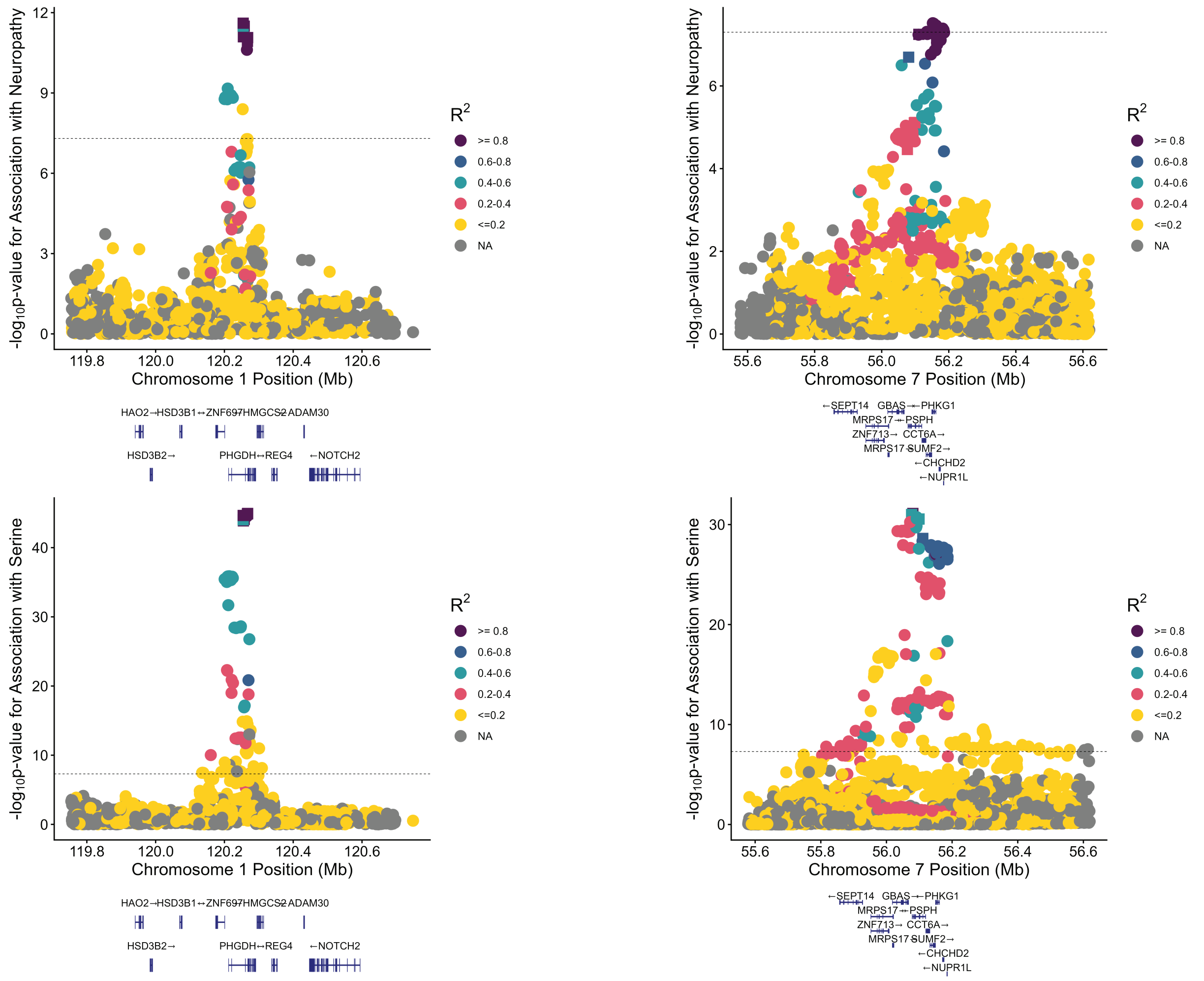


**A.**

| Colocalization results | |
|---|---|
| Hypothesis | Posterior probability of hypothesis |
| $H_0$ | 2.7e-47 |
| $H_1$ | 1.5e-41 |
| $H_2$ | 1.6e-8 |
| $H_3$ | 7.8e-3 |
| $H_4$ | 0.992 |

| 95% Credible Set | |
|---|---|
| Variant | Posterior probability of causality |
| rs562038 | 0.33 |
| rs483180 | 0.29 |
| rs662602 | 0.14 |
| rs477992 | 0.09 |
| rs561931 | 0.08 |
| rs478093 | 0.03 |

**B.**

| Colocalization results | |
|---|---|
| Hypothesis | Posterior probability of hypothesis |
| $H_0$ | 4.7e-29 |
| $H_1$ | 1.3e-26 |
| $H_2$ | 1.0e-3 |
| $H_3$ | 0.27 |
| $H_4$ | 0.72 |

| 95% Credible Set | |
|---|---|
| Variant | Posterior probability of causality |
| rs62457261 | 0.93 |
| rs2016724 | 0.008 |
| rs6955423 | 0.006 |
| rs6951810 | 0.006 |
| rs4948102 | 0.006 |

**Supplemental Figure 7**. Bayesian colocalization analysis evaluating overlap of GWAS associations with neuropathy in individuals with diabetes and with plasma serine levels in Lotta et al [REF] at two loci associated with both traits at genome-wide significance, *PHGDH* **(A)** and *PSPH* **(B)**. Plots show SNP associations with neuropathy (top) and with serine levels (bottom); in each plot, color corresponds to linkage disequilibrium (LD) with the lowest p-value SNP at the locus for each trait, and square-shaped SNPs are members of the 95% credible set of variants based on posterior probability for causality for both neuropathy and serine levels at the locus. Colocalization results for each locus are quantified as the posterior probability in support of five hypotheses: $H_0$ that the data do not support a causal genetic association with either trait; $H_1$ that the data support a causal genetic association for trait 1 (neuropathy) but not trait 2 (serine levels); $H_2$ that the data support a causal genetic association for trait 2 (serine) but not trait 1 (neuropathy); $H_3$ that the data support a causal genetic association for both traits but without a shared causal variant; and $H_4$ that the data support a shared causal genetic variant for both traits. For each SNP analyzed, *coloc* also provides the posterior probability that the variant is the shared causal variant for both traits assuming $H_4$ is true, and the "credible set" of variants whose posterior probabilities sum to >0.95 are shown for each locus.

### Supplemental Figure 8.

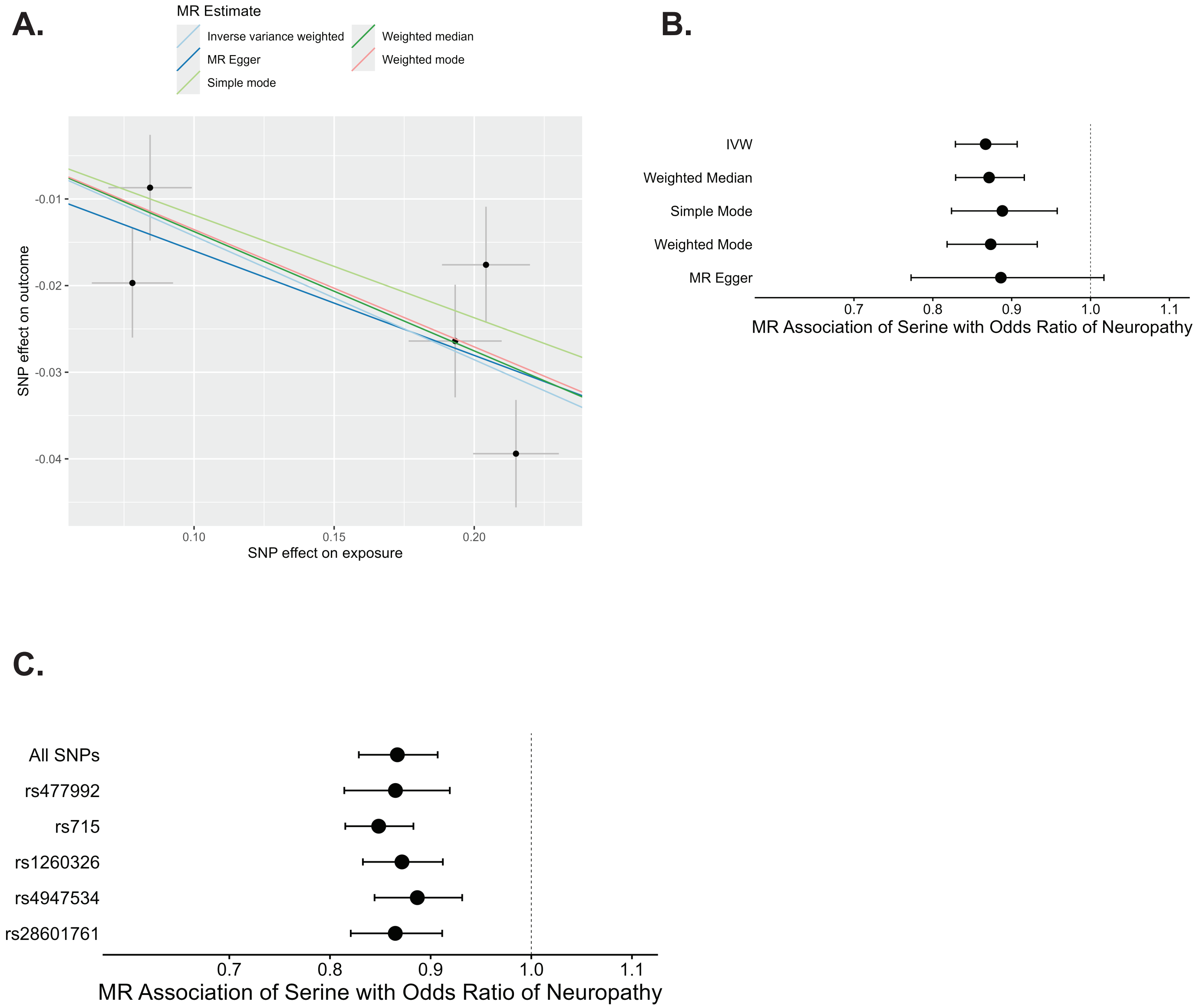
